# Automated Monitoring of Clinical Practice Guideline Adherence Using FHIR and OMOP: A Multi-Center Study in Intensive Care Units

**DOI:** 10.1101/2025.01.02.24319597

**Authors:** Gregor Lichtner, Fridtjof Schiefenhövel, Bora Gashi, Ingrid Martin, Carlo Jurth, Lisa Vasiljewa, Dana Kleimeier, Sebastian Gibb, Markus Heim, Jürgen Brugger, Johannes Lohr, Martin A. Feig, Saya Speidel, Thomas Bienert, Igor Abramovich, Mathias Kaspar, Anja Sindel, Laurenz Mehringer, Ludwig Christian Hinske, Philipp Simon, Axel R Heller, Peter Kranke, Patrick Meybohm, Felix Balzer, Claudia Spies, Gerhard Schneider, Klaus Hahnenkamp, Dagmar Waltemath, Martin Boeker, Falk von Dincklage

## Abstract

**Background:** Clinical practice guidelines are important tools for clinical decision support, but monitoring guideline adherence manually is highly resource-intensive. Therefore, we developed an automated system for evaluating guideline adherence based on computer-interpretable representations of guidelines. We implemented the system across multiple university hospitals and assessed its validity and performance by comparing its guideline adherence evaluations to those conducted by medical professionals.

**Methods:** We selected six representative clinical guideline recommendations from across 41 intensive care guidelines and translated these text-based recommendations into a computer-interpretable, Fast Healthcare Interoperability Resources (FHIR)-based format using an iterative consensus approach. Clinical data from five university hospitals were transformed into the Observational Medical Outcomes Partnership (OMOP) common data model. A decision support system was developed to interpret FHIR-encoded recommendations and apply them to OMOP-formatted patient data. We evaluated the system retrospectively on intensive care data covering 3.5 years and validated its performance by comparing system-generated decisions with human decisions in three hospitals. We created and iteratively refined a user interface for individual and ward-level adherence visualization.

**Findings:** We expert-reviewed more than 18,000 patient days to assess the applicability of and adherence to the recommendations. The system demonstrated 97.0% accuracy in identifying guideline applicability and adherence, with significantly higher accuracy than human reviewers (accuracy 86.6%, p<0.001, McNemar’s Test). The automated system processed more than 2000 patient days per second for a total of 2,200,000 patient days across 82,000 intensive care episodes, compared to humans’ two patient days per minute.

**Interpretation:** We demonstrate that an automated adherence monitoring system outperforms human reviewers in both accuracy and time efficiency. Using FHIR-encoded recommendations enables flexibility and scalability across hospitals with different data infrastructures. Future work should focus on integrating unstructured patient data and expanding the range of encoded recommendations.

**Funding:** Federal Ministry of Education and Research of Germany.

## 1 Introduction

Clinical practice guidelines are systematically developed statements designed to guide health care professionals and patients in evidence-based decision-making.^1^ Adherence to guidelines ensures that patients receive standardized care, leading to improved patient outcomes and reduced health care costs.^2–4^ However, several challenges hinder the implementation of guidelines in clinical practice, including a lack of awareness of guideline contents, uncertainties regarding their applicability in specific clinical circumstances, as well as a lack of technology to automatically integrate guidelines in care practice.^5^ These challenges are particularly pronounced in intensive care units, where patient conditions are complex and constantly changing, and treatment often involves multiple medical specialties.^6–8^ The high complexity and volume of patient data in intensive care settings can make it difficult for physicians to accurately assess guideline adherence consistently.

Automated systems using computer-interpretable guidelines have the potential to significantly enhance guideline adherence monitoring^9^ and to improve overall quality of care.^10,11^ In contrast to systems that are based on system-integrated encodings of guideline recommendation rulesets, systems processing externally provided computer-interpretable guidelines have a higher flexibility to adapt to additional guidelines or updated recommendations.^9^ To employ guidelines in such systems, they need to be represented in a computer-interpretable format, for which a wide range of representation formalisms have been developed.^12–14^ While these formalisms have proven to be effective, they often require adaptation to local data models, limiting their scalability and broader applicability. To address this, we have previously developed a Fast Healthcare Interoperability Resources (FHIR) implementation guide – one of the most important standards for representing and exchanging health care information – for the computer-interpretable representation of clinical practice guidelines including the underlying evidence used to create these guidelines.^15^ FHIR is particularly well-suited for the purpose of this work due to its ability to standardize data exchange across health care systems.

One significant barrier to scalability of automated monitoring solutions is the requirement of adapting computer-interpretable guidelines to local data models used by different healthcare providers. To address this challenge of differences in local data models, the Observational Medical Outcomes Partnership (OMOP) Common Data Model (CDM) has emerged as a widely adopted standard for representing clinical data in a consistent format.^16^ The OMOP CDM has been implemented in hospitals and healthcare systems to improve data interoperability and enable more effective use of clinical data for research and decision support.^17,18^

The aim of this study was to evaluate the accuracy and efficiency of a novel automated system that combines FHIR for computer-interpretable guideline representation and the OMOP CDM for patient data to monitor adherence to clinical practice guidelines in intensive care settings. By leveraging these interoperable standards, the system is designed to be adaptable across different healthcare environments, addressing the challenges of scalability and consistency that have limited previous approaches. To validate its robustness and generalizability across diverse clinical settings, we implemented the system in multiple university hospitals and compared its performance against human experts.

## 2 Methods

### 2.1 Selection of Clinical Practice Guideline Recommendations

To select representative guideline recommendations for integration in our system, we conducted a multi-step process involving relevance and suitability ratings by clinical experts. From 41 intensive care guidelines with a total of 1,597 recommendations, we ultimately selected six recommendations based on expert ratings, focusing primarily on COVID-19-related treatments to demonstrate the system’s adaptability to rapidly evolving guidelines (**Table 1**). Details of the selection process are provided in the Appendix.

**Table 1:** Overview of Clinical Recommendations Implemented for Automated Adherence Monitoring. Shown are the six clinical guideline recommendations that were selected and implemented for automated monitoring using a FHIR-based format. “No” refers to the original numbering of the recommendation within the respective guideline. “Guideline” specifies the source document from which the recommendation was derived. The “Recommendation” column shows the machine-translated content of each recommendation, which outlines specific clinical interventions related to COVID-19 and ARDS management. ARDS: Acute Respiratory Distress Syndrome; COVID-19: Coronavirus Disease 2019; PaO_2_: Partial pressure of arterial oxygen; FiO_2_: Fraction of inspired oxygen; PEEP: Positive end-expiratory pressure; VT: tidal volume.

| No | Short Name | Guideline | Recommendation |
| --- | --- | --- | --- |
| 15 | Prophylactic Anticoagulation | S3 guideline Recommendations for the inpatient treatment of patients with COVID-19 - Living Guideline <sup>19</sup> | Hospitalized patients with COVID-19 should receive standard drug thromboembolism prophylaxis with low molecular weight heparin in the absence of contraindications. Alternatively, fondaparinux may be used. |
| 18 | No Therapeutic Anticoagulation |  | Therapeutic anticoagulation should not be used in intensive care patients without a specific indication (e.g., pulmonary embolism). |
| 35 | Tidal Volume | | In ventilated patients with COVID-19 and ARDS, tidal volume should be $\leq 6$ ml/kg standard body weight, end-inspiratory airway pressure $\leq 30$ cm H<sub>2</sub>O. |
| 36a | PEEP |  | For the orientational adjustment of PEEP in COVID-19, the FiO<sub>2</sub>/PEEP table of the ARDS Network should be considered. Close monitoring can adjust the PEEP to the individual patient's situation. |
| 36b | Prone Positioning | | In ARDS and at PaO<sub>2</sub>/FiO<sub>2</sub> $< 150$ mmHg, abdominal positioning should be consistently performed, with an abdominal positioning interval of at least 16 hours. |
| M.1 | Sepsis Tidal Volume | Sepsis - prevention, diagnosis, therapy and follow-up care <sup>20</sup> | We recommend ventilating patients with ARDS with a VT $\leq 6$ ml/kg ideal body weight. |

### 2.2 Encoding Recommendations Using FHIR

Recommendations were encoded in FHIR following a structured process based on our previous work.^21^ In short, an intensive care physician and a medical computer scientist jointly decomposed the recommendations and extracted and mapped relevant medical concepts to standardized terminologies. The recommendations were then encoded in FHIR according to the Clinical Practice Guidelines (CPG)-on-Evidence-based Medicine (EBM)onFHIR Implementation guide.^15^ The encoded recommendations were syntactically and semantically validated and reviewed by multiple intensive care specialists and medical computer scientists to ensure accuracy. Further details of the encoding process are provided in the Appendix.

### 2.3 Data Transformation to the OMOP CDM

As different hospitals have different underlying data models in their intensive care information systems, we chose to map all local concepts onto the OMOP CDM as the standardized data model for our guideline-adherence monitoring system. Each of the five hospitals employs a unique combination of information systems, from a total of 5 different vendors (for details see appendix). We developed *extract, transform, load* (ETL) pipelines for each hospital individually to transform the necessary data to the OMOP CDM. To avoid ambiguities, we provided a conversion guide to all participating hospitals, detailing the expected ways of representing the required data. After conversion, the data distributions of all transformed variables were manually checked for any deviations from expected distributions.

### 2.4 Development of the Execution Engine

The execution engine is the central software component of the guideline-based decision support system (**Figure 1**). It ingests clinical practice guideline recommendations in CPG-on-EBMonFHIR format and interprets them to create appropriate queries against an OMOP CDM database, which it then combines according to the logic specified in the CPG-on-EBMonFHIR-based recommendations to compute the individual applicability of and adherence to the recommendations. The system follows a modular architecture,^9^ where the execution engine retrieves the computer-interpretable recommendations from an external FHIR server, interprets them and executes SQL statements against an external relational patient database implementing the OMOP CDM. For more details on the execution of guidelines, see the Appendix.

**Figure 1:**
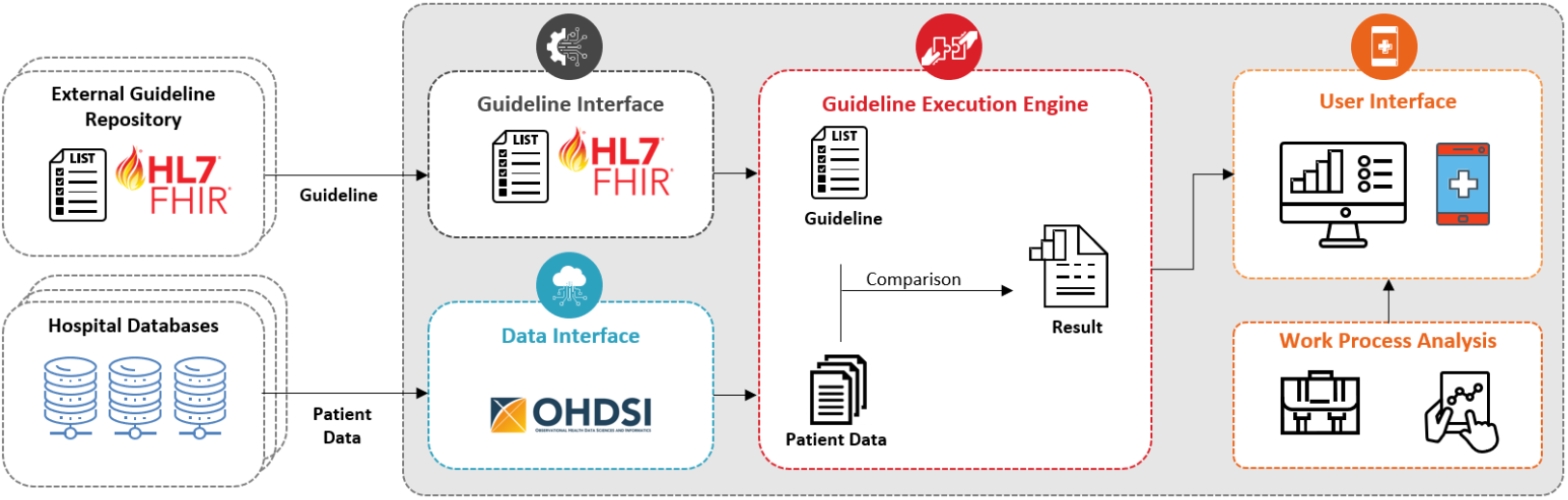
Architecture of the Developed Guideline-Based Decision Support System. Shown is the architecture of the system for monitoring guideline adherence. FHIR-encoded guidelines are retrieved from external repositories via the Guideline Interface, while patient data from clinical databases, standardized to the OMOP common data model, are accessed through the Data Interface. The Guideline Execution Engine interprets the computer-interpretable guideline recommendations and ingests the patient data required to evaluate the applicability and adherence to the recommendations. Results can be displayed on an exchangeable User Interface, which can be adapted to specific workflows and requirements for real-time or retrospective guideline adherence monitoring.

### 2.5 Evaluation of the Automated Guideline Recommendation Adherence

To evaluate for each patient and each recommendation whether the recommendation is applicable and whether the patient is treated accordingly, we ran the execution engine in all five participating hospitals on data from January 2020 to May 2023, covering the entire COVID-19 pandemic in Germany. The execution engine divides the time span for each patient into mutually exclusive time intervals to indicate whether the patient meets the population criteria (i.e., whether the patient is part of the recommendation’s target group), the intervention criteria (i.e., whether the patient is treated according to the recommendation), and the combination of both. These time intervals are classified as *positive* if the criteria are fulfilled, *negative* if they are not, or *no-data* if insufficient data is available to decide. For the combination of population and intervention, time intervals may also be classified as *not-applicable*, indicating that the recommendation does not apply to the patient during that period.

For determination of daily adherence, a patient is considered part of the recommendation’s population if the population criteria are met at any point during the day, even if not for the entire day. Adherence to the recommendation is considered met if, on that day, the criteria for both population and intervention are fulfilled, or the recommendation is marked as *not-applicable* for the patient. However, if the recommendation is applicable but not followed at any point during the day, the whole day is classified as non-adherent for the patient.

### 2.6 Validation Study

To validate the transformation process from the text-based guideline recommendations to the computer-interpretable recommendations and the technical correctness of the execution engine, we performed a validation study in three of the participating hospitals. To that end, we selected ten days, each spaced apart at least 60 days to minimize statistical bias (for details see Appendix).

In the three hospitals participating in the validation study, we recruited a total of seven intensive care physicians that were not involved otherwise in the study and had no information about the implementation of the computer-interpretable recommendations or the software. All participating physicians had at least one year of intensive care experience and were proficient in the patient data management system used in their respective hospitals. Each clinician was provided the text-based, original version of each of the six recommendations, and they were instructed to manually check for each intensive care patient that was present on the specified days and for each of the six recommendations, whether the recommendations were applicable to the patient (i.e. if the patient was part of the population) on the selected day. If this was the case, they further checked whether the patient was treated according to the recommendation. The clinicians were instructed to base their answer on the patient data available from the clinical information systems, but without instructions regarding which data they should look at or how to interpret the text-based guideline recommendation.

Each clinician reviewed different days, but to compare their ratings and determine the inter-rater-reliability, in each clinic we assigned one or two of the days to all raters. These were the dates with the highest number of patients treated according to the recommendations, as output by the execution engine.

To compare the performance of the system against that of the human reviewers, in one of the validation sites, we additionally recruited two clinically highly experienced supervising physicians to thoroughly re-check all cases with disagreement between the raters and the execution engine and thereby produce the *ground truth* of ratings against which the performance of the system as well as the performance of the raters could be quantified with. Therefore, to quantify performance of the system and the human raters, their ratings were compared against those of the *ground truth* by the experts and performance metrics (sensitivity, specificity, accuracy, precision and F1 score) were calculated.

### 2.7 Statistical Analysis

We used McNemar’s test as implemented in the Python *mlxtend* package to compare the classification accuracy of the system against a human rater, with the expert annotations as the ground truth. A 2×2 contingency table was constructed, and the test was applied with a continuity correction to assess performance differences.

### 2.8 Ethics Approval

This study was approved by the local ethics committees of the participating hospitals (for details see Appendix).

## 3 Results

### 3.1 Patient Cohort

We converted the clinical data from five university hospitals between January 1, 2020 and May 31, 2023 from the respective intensive care data systems to the OMOP CDM, for a total of 82,015 individual intensive care episodes (Supplementary Tables 2-5).

### 3.2 Validation Findings

Overall, the system achieved a 97.0% accuracy in identifying guideline applicability and adherence. This performance significantly surpassed that of human reviewers, who achieved an accuracy of 86.6% (McNemar’s Test; χ^2^ = 149.93; p < 0.001) (**Table 2**). Both the system and the human raters correctly classified 1,884 cases (83.9%). The system alone misclassified 63 cases (2.8%) that the human raters got right, while the human raters misclassified 296 cases that the system got right (13.2%). Both the system and the human raters incorrectly classified 4 cases (0.2%).

**Table 2:** Performance Metrics of System-Generated and Physician-Reviewed Decisions vs the Ground Truth Decisions, Determined by Experienced Supervising Physicians.

|  | System vs Ground Truth | Physicians vs Ground Truth |
| --- | --- | --- |
| Accuracy | 97.0% | 86.6% |
| Sensitivity | 81.4% | 80.8% |
| Specificity | 99.5% | 87.6% |
| Precision | 96.6% | 51.2% |
| F1 | 88.3% | 62.7% |
| Prevalence | 13.9% | 13.9% |
| Total N | 2247 | 2247 |

Regarding time efficiency, the human review of 18,115 patient days (calculated as patients × days × recommendations) required a total duration of 150 hours across all three hospitals, which is approximately 30 sec per patient, day and recommendation. In contrast, the system processing allowed evaluation of >2000 patient days per second on a standard computer.

### 3.3 Sources for Mismatches Between System and Raters

We categorized all mismatches between the system and the physician raters into rater errors, where the system’s decision was actually correct, or system errors, where the raters’ decision was actually correct (**Figure 2**). For the system errors, further analysis revealed that they almost exclusively arose from information contained in the primary database as unstructured free-text fields, which were not transferred through the ETL pipelines into the OMOP CDM and were therefore unavailable to the system. In contrast, human raters, who accessed the primary information systems directly through their user interfaces, could review both structured and unstructured data, accounting for the observed discrepancies.

**Figure 2:**
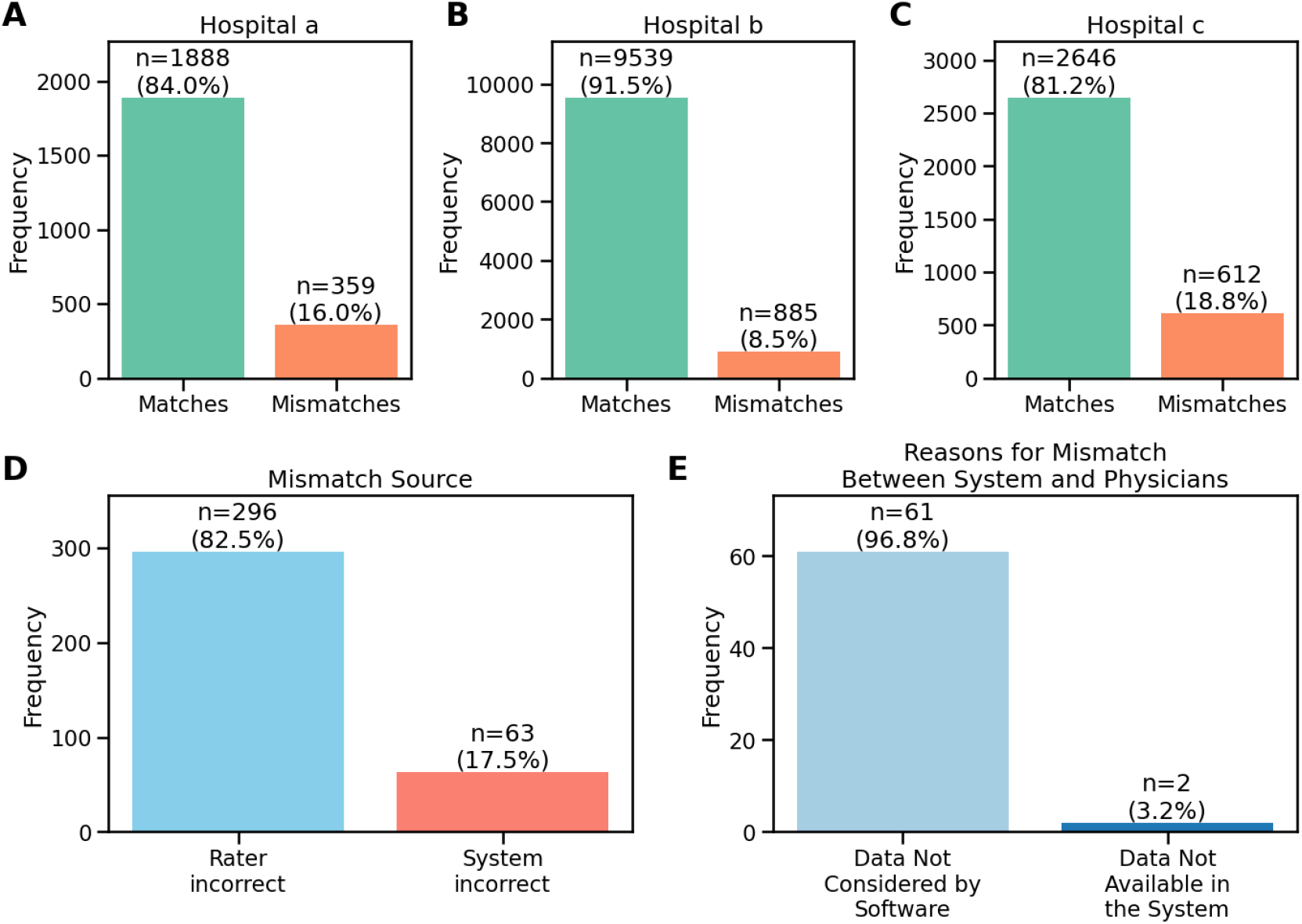
Comparison of System-Generated Decisions with Physician Reviews and Analysis of Mismatches. Shown are the agreement rates between system-generated decisions and physician - reviewed decisions for three hospitals (**A-C**), the source of the mismatches for hospital a, indicating the number of cases where the physician reviewers were incorrect and cases where the system was incorrect, as determined by supervising physicians during a second review (**D**), and a breakdown of the primary sources of system errors (**E**).

### 3.4 Guideline Recommendation Adherence

Automated evaluation of guideline adherence showed a diverse picture, where the relative adherence to the recommendations depends both on the recommendation and the hospital (**Figure 3**).

**Figure 3:**
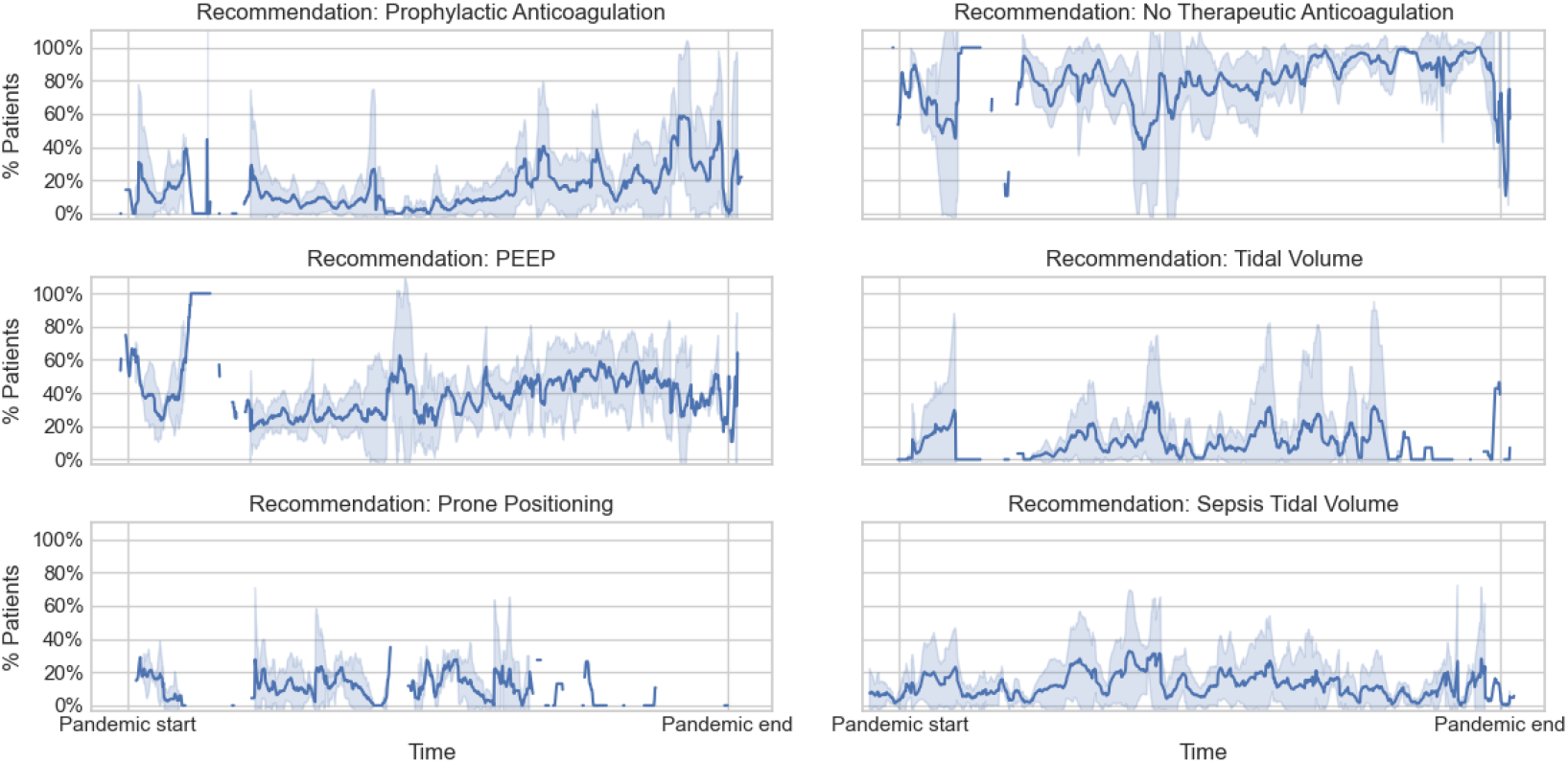
Adherence Rates Across Recommendations and Hospitals. Shown are the mean adherence rates for the five hospitals for each of the six recommendations across the whole COVID-19 pandemic, showing how adherence to the recommendation evolved over time. The percentage is calculated as the number of patients meeting the recommendation’s criteria and receiving the recommended treatment, divided by the number of patients meeting the recommendation’s criteria. The adherence for days without any patients to which the recommendation was applicable is left blank in the plots. Shaded area: 95% confidence interval.

## 4 Discussion

In this study, we developed an automated guideline adherence monitoring system that leverages FHIR-encoded, computer-interpretable recommendations and applies them to OMOP CDM-encoded patient data to determine individual guideline adherence. We deployed the system across five university hospitals with unique IT infrastructures to assess the adherence to six recommendations across over 82,000 intensive care episodes. By establishing a ground truth through manual chart review by highly experienced supervising physicians, we demonstrate that the system significantly outperforms human raters in both accuracy and time efficiency.

### 4.1 Errors in Human versus Automated Interpretation of Guideline Recommendations

We found that over 80% of discrepancies between the system and the human raters were due to human error rather than system errors. A major source of human error is the overlooking of information during review of large volumes of data in the frontends of critical care information systems (CCIS). For instance, frequently recorded values, such as parameters from mechanical ventilation, may be difficult to assess repeatedly for human reviewers, leading to values being overlooked. In some frontends of CCIS, repeated values may also be hidden if the scaling of data is not adjusted properly by the user. This makes it particularly difficult for humans to comprehensively review all values.

Another factor is the inconsistent interpretation of guideline recommendations by different human raters, especially when recommendations are imprecisely formulated or do not cover all required details and instead rely on implicit knowledge. For example, the *Tidal Volume* recommendation does not provide the formula for calculating the ideal body weight. Depending on the knowledge of the human rater, different formulas for calculating the ideal body weight might be used. Similarly, in the context of drug-related actions, recommendations often refer to contraindications, without clearly defining these and thus requiring human raters to rely on their own understanding, which can vary in detail.

System errors were far less frequent than human errors (**Figure 2** D), and the large majority of the system errors were related to data not being considered by the software, often because diagnoses were recorded in free-text fields such as patient history notes, rather than being coded in a structured way. Despite these limitations, system errors were relatively rare compared to human errors and, importantly, the system is highly scalable. Unlike human reviewers, who face challenges in reviewing large amounts of data and maintaining consistency,^22^ the system can efficiently process large datasets with high accuracy.

### 4.2 Development of the Execution Engine to Monitor Clinical Practice Guideline Adherence using FHIR-encoded Guidelines

The execution engine that we developed and validated in this study provides a flexible and adaptable framework for automated guideline adherence monitoring of FHIR-encoded clinical practice guidelines. While the primary use case in this study involved FHIR-encoded guidelines, the engine’s structure allows recommendations to be defined programmatically, enabling adaptability to various guideline formats. By testing the system across five university hospitals, each with its unique set of clinical information systems and underlying data models, we demonstrated its robustness in diverse clinical environments. The use of interoperable data and guideline representation formats significantly enhances the generalizability of our approach. A key feature of our system is the specification of recommendations in a format according to level 3 of Boxwala’s model of knowledge representation,^23^ rather than specifying the recommendations as computer-executable code (level 4 representation). This allows the system to integrate additional guidelines without the need for additional software development.

Automated guideline-adherence monitoring systems hold significant potential to improve quality management in clinical practice by identifying areas where guideline adherence is lacking and by providing individualized clinical decision support. For such a system to gain acceptance by the healthcare personnel, two key factors are essential: usability and trust.^24,25^ One aspect to high usability is the integration of the system into the clinical workflow in a way that prevents disruption of the care processes. To ensure this, we conducted a work process analysis of guideline adherence monitoring in the intensive care setting. We used insights gained from that analysis to design a user interface that aligns with the needs of quality management in critical care (see Appendix). An iterative design approach, which involved multiple rounds of feedback from intensive care physicians, allowed us to refine the user interface. Additionally, the adherence analysis returned by the execution engine can be integrated either directly into existing clinical information systems or into standalone user interfaces optimized for specific tasks, providing flexibility to suit different clinical and user demands.

The second critical factor for a clinical decision support system to gain acceptance among healthcare personnel is trust in the system’s decisions. By using explicit and deterministic rules that are transparently derived from evidence-based guidelines, the system’s decisions are traceable and justifiable. To support this transparency, we included in our user interface the ability to examine the underlying patient data that the system uses to come to its decision whether a recommendation is applicable and adhered to, promoting trust in the system’s output.

### 4.3 Factors Influencing Clinical Practice Guideline Adherence

Analysis of the implemented recommendations revealed relatively low adherence rates, mostly between 10-60%, except for one “negative” recommendation that is naturally fulfilled by the absence of an action. Even though achieving 100% adherence is unrealistic due to justified deviations from recommended treatments for medical reasons, our reported adherence rates of 10-60% are relatively low compared to previous studies.^9,26–29^

Several factors contributed to the low observed adherence rates. First, we here focused on COVID-19-related recommendations over the course of the pandemic, during which these recommendations were just newly introduced. Therefore, limited awareness and initial uncertainty regarding their reliability likely contributed to low adherence, especially in the early phases. Accordingly, adherence increased over time, as observed for recommendations on anticoagulatory treatment and prone positioning of COIVD-19 patients.

Second, incomplete documentation can lead to apparent non-adherence. For example, inconsistent documentation of prone positioning timing in some hospitals made it difficult to assess whether patients were placed in prone position according to the recommended duration, possibly leading to not properly detected adherence.

Third, guideline recommendations are not typically developed with the intent of enabling rigid adherence monitoring. Accordingly, studies often evaluate guideline adherence less strictly than the exact specification of the recommendation, as this would not align with clinical judgement of guideline-adherent patient care. However, any deviation from the explicit specification reduces the reproducibility and comparability of the adherence monitoring. For example, a strict evaluation classifies a patient’s treatment as non-adherent to the *Tidal Volume* recommendation if the tidal volume briefly exceeds the threshold defined in the guideline, which, while technically accurate, is not clinically meaningful. It therefore seems reasonable to define permissible deviation rates to ensure that patient treatment can still be considered guideline-adherent despite occasional deviations.

Similarly, imprecisely defined recommendations relying on implicit knowledge can lead to variability. For example, drug-related recommendations require complete clarity regarding permissible drugs, dosages and administration frequency. Furthermore, timing considerations need to be specified when multiple different data points are evaluated, such as the acceptable time difference between FiO_2_ and PEEP measurements to be considered related.

Therefore, to ensure reproducible and comparable guideline adherence monitoring, it is in our view advisable to consider adherence evaluation already during the development of guideline recommendations This approach, which is already being recommended through initiatives to co-develop human-readable and computer-interpretable guidelines, would ensure the necessary specificity, completeness and defined ranges for permissible deviations.^5,30^ To ensure comparable evaluations of guideline adherence, we recommend that guidelines explicitly define: (i) tolerance ranges for target values and dosages, (ii) requirements for frequency or duration of adherence (e.g., whether conditions must be met continuously or at specific intervals), (iii) acceptable limits for the frequency of deviations from recommended values, and (iv) defined time intervals within which measurements must be recorded to be considered relevant to the same adherence evaluation.

### 4.4 Limitations

One limitation of our study was that the system validation was performed retrospectively, without evaluating its use for quality management or decision support prospectively. However, assessing guideline adherence using the here developed system is a deterministic process and produces consistent outcomes regardless of the time of evaluation.

Furthermore, there is potential for bias in the expert assessments that were used as ground truth, which could have influenced the validation outcomes. To address this, we have recruited two independent supervising physicians for ground truth labeling in order to reduce bias.

As a potential limitation of external validity of our results, the scope of our study was restricted to a handful of guideline recommendations, and the accuracy of the system is dependent on the accessibility and quality of the data required for assessing adherence to these recommendations. To address the variability regarding these requirements, we aimed to select recommendations that cover different aspects of intensive care with different requirements on the availability and quality of the data.

### 4.5 Conclusion

This study demonstrates the feasibility and effectiveness of an automated system for monitoring guideline adherence in intensive care settings. Our approach, combining FHIR-encoded clinical guidelines with OMOP-formatted patient data, significantly improves both the accuracy and efficiency of guideline adherence monitoring compared to human reviewers. The system provides a scalable and reliable method for monitoring guideline adherence across different healthcare institutions and a foundation for further development towards broader integration of evidence-based medicine in clinical practice.

## Data Availability

All code developed in this project has been published under https://github.com/CODEX-CELIDA/. The CPG-on-EBMonFHIR-encoded recommendations have been made available under https://github.com/CODEX-CELIDA/celida-recommendations/releases.
Individual patient data cannot be shared due to data protection regulations. Anonymized (e.g. aggregated) data can be shared upon reasonable request to the corresponding author.

https://github.com/CODEX-CELIDA/

## 6 Acknowledgements

We thank Burkhard Meißner and Matthäus Morhart for their support with data conversion and integration.

## 8 Contributors

Conceptualization: GL, PM, CS, GS, DW, MB, FvD. Data Curation: GL, FS, BG, IM, CJ, DK. Formal Analysis: GL, FvD. Funding Acquisition: PM, FB, CS, GS, DW, MB, FvD. Investigation: GL, FS, BG, IM, CJ, LV, DK, SG, MH, JB, JL, MF, SS, TB, IA, MK, AS, LM, PK, FvD. Methodology: GL, DW, MB, FvD. Project Administration: GL. Resources: GL, MK, LH, PS, ARH, PK, PM, FB, CS, GS, KH, DW, MB, FvD. Software: GL, FS, BG, IM, DK. Supervision: GL, MK, LH, PS, ARH, PK, PM, FB, CS, GS, KH, DW, MB, FvD. Validation: GL, FS, BG, IM, CJ, SG, JL, MF, SS, TB, IA, MK, AS, LM, FvD. Visualization: GL. Writing – Original Draft: GL, FvD. Writing – Review & Editing: GL, FS, BG, IM, CJ, LV, DK, SG, MH, JB, JL, MF, SS, TB, IA, MK, AS, LM, LH, PS, ARH, PK, PM, FB, CS, GS, KH, DW, MB, FvD. All authors revised and edited the manuscript critically for important intellectual content. All authors had final responsibility for the decision to submit for publication.

## 9 Data sharing

All code developed in this project has been published under https://github.com/CODEX-CELIDA/. The CPG-on-EBMonFHIR-encoded recommendations have been made available under https://github.com/CODEX-CELIDA/celida-recommendations/releases.

Individual patient data cannot be shared due to data protection regulations. Anonymized (e.g. aggregated) data can be shared upon reasonable request to the corresponding author.

