## Appendix for "Automated Monitoring of Clinical Practice Guideline Adherence Using FHIR and OMOP: A Multi-Center Study in Intensive Care Units"

### 1 Methods

#### 1.1 Selection of Clinical Practice Guideline Recommendations for Integration

To select representative guideline recommendations for integration into the guideline-based decision support system, we conducted a multi-step process. First, we identified clinical practice guidelines for intensive care through searches in the German guideline repository of the "Arbeitsgemeinschaft Wissenschaftlicher Medizinischer Fachgesellschaften" (AWMF), the guidelines international network and relevant organizational websites (European Resuscitation Council (ERC), European Society of Intensive Care Medicine (ESICM), and European Society of Anaesthesiology and Intensive Care (ESAIC)). Searches were conducted in January 2022 and 41 guidelines published by recognized intensive care organizations, focusing on intensive care and available in full-text, were included (**Supplementary Table 1**). From these guidelines, a total of 1,597 recommendations were extracted. Five raters independently rated all recommendations regarding clinical relevance using a three-point scale (high relevance, moderate relevance, or low relevance). Recommendations rated as highly relevant by at least three of five raters (n=547) were subjected to a second round of relevance rating using the same scheme, aiming to reduce the recommendations to no more than one-third of the total. This process resulted in 206 recommendations from 26 guidelines.

These 206 recommendations were then rated by seven certified intensive care specialists for their suitability for integration into the guideline-based decision support system on a scale from 0 (not suitable) to 10 (perfectly suitable). Based on recommendations with the highest average ratings, we selected five COVID-19-related treatment recommendations that covered four aspects of treatment that each refer to different data domains (medication, laboratory values, ventilation and positioning) and a sixth non-COVID-19-related treatment. We focused on COVID-19-related treatment recommendations to demonstrate the system’s utility in rapidly changing recommendations (as part of a living guideline). This allowed us to analyze a possible time-dependence of implementation of these recommendations during the course of the pandemic.

#### 1.2 Guidelines Included in Relevance Screening

| No | Title | AWMF Register No | Version | Ref |
|---|---|---|---|---|
| **1** | S3-Leitlinie Analgesie, Sedierung und Delirmanagement in der Intensivmedizin (DAS-Leitlinie 2015) | 001-012 | 2015 | [1] |

| No | Title | AWMF Register No | Version | Ref |
|---|---|---|---|---|
| 2 | S2e-Leitlinie – „Lagerungstherapie und Frühmobilisation zur Prophylaxe oder Therapie von pulmonalen Funktionsstörungen“ | 001-015 | 2015 | [2–4] |
| 3 | S2k-Leitlinie Klinische Ernährung in der Intensivmedizin | 073-004 | 2.0 from 06/2018 | [5, 6] |
| 4 | S2k-Leitlinie Behandlung der Thorakalen Aortendissektion Typ A | 011-018 | 1.1 from 02/2021 | [7, 8] |
| 5 | S2k-Leitlinie Gastrointestinale Blutung | 021-028 | 1.0 from 05/2017 | [9, 10] |
| 6 | S2k-Leitlinie Venenthrombose und Lungenembolie: Diagnostik und Therapie | 065-002 | 2015 | [11] |
| 7 | S2k-Leitlinie Prolongiertes Weaning | 020 - 015 | 2.0 from 08/2019 | [12, 13] |
| 8 | S2k-Leitlinie Behandlung von spontanen intrazerebralen Blutungen | 030 - 002 | 6.2 from 04/2021 | [14] |
| 9 | S2k-Leitlinie Akute Herzinsuffizienz und mechanische Kreislaufunterstützung | 023 - 032 | 2.0 from 02/2020 | [15] |
| 10 | S2k-Leitlinie Komplikationen der Leberzirrhose | 021 - 017 | 2.1 from 11/2018 | [16, 17] |
| 11 | S3-Leitlinie Sepsis - Prävention, Diagnose, Therapie und Nachsorge | 079 - 001 | 3.1 from 12/2018 | [18, 19] |
| 12 | S3-Leitlinie Intravasale Volumentherapie beim Erwachsenen | 001 - 020 | 2.0 from 07/2020 | [20, 21] |
| 13 | S3-Leitlinie Sauerstoff in der Akuttherapie beim Erwachsenen | 020 - 021 | 1.0 from 06/2021 | [22–24] |
| 14 | S3-Leitlinie Epidemiologie, Diagnostik und Therapie erwachsener Patienten mit nosokomialer Pneumonie – Update 2017 | 020 - 013 | 2017 | [25, 26] |
| 15 | S3-Leitlinie Extrakorporale Zirkulation (ECLS / ECMO), Einsatz bei Herz-und Kreislaufversagen | 011 - 021 | 1.0 from 08/2020 | [27, 28] |
| 16 | S3-Leitlinie Prophylaxe der venösen Thromboembolie (VTE) | 003 - 001 | 3.0 from 10/2025 | [29, 30] |
| 17 | S3-Leitlinie Invasive Beatmung und Einsatz extrakorporaler Verfahren bei akuter respiratorischer Insuffizienz | 001 - 021 | 1.0 from 11/2017 | [31] |
| 18 | S3-Leitlinie Infarkt-bedingter kardiogener Schock - Diagnose, Monitoring und Therapie | 019 - 013 | 2.0 from 02/2019 | [32, 33] |
| 19 | S3-Leitlinie Empfehlungen zur stationären Therapie von Patienten mit COVID-19 - Living Guideline | 113 - 001 | 10/2021 | [34, 35] |
| 20 | S3-Leitlinie Polytrauma / Schwerverletzten-Behandlung | 187 - 023 | 07/2016 | [36] |
| 21 | S1-Leitlinie Neurogene Dysphagie | 030 - 111 | 4.0 from 02/2020 | [37] |
| 22 | S1-Leitlinie Hypoxisch-ischämische Enzephalopathie (HIE) im Erwachsenenalter | 030 - 119 | 09/2017 | [38] |
| 23 | S1-Leitlinie Intrakranieller Druck (ICP) | 030 - 105 | 09/2017 | [39] |
| 24 | European Resuscitation Council Guidelines 2021: Adult advanced life support | | 2021 | [40] |
| 25 | European Resuscitation Council and European Society of Intensive Care Medicine Guidelines 2021: Post-resuscitation care | | 2021 | [41] |
| 26 | ERC-ESICM guidelines on temperature control after cardiac arrest in adults | | 2021 | [42] |
| 27 | European Society of Clinical Microbiology and Infectious Diseases (ESCMID) guidelines for the treatment of infections caused by multidrug-resistant Gram-negative bacilli (endorsed by European society of intensive care medicine) | | 2022 | [43] |
| 28 | Transfusion strategies in non-bleeding critically ill adults: a clinical practice guideline from the European Society of Intensive Care Medicine | | | [44] |
| 29 | Noninvasive respiratory support in the hypoxaemic peri-operative/periprocedural patient: a joint ESA/ESICM guideline | | | [45] |

| No | Title | AWMF Register No | Version | Ref |
|---|---|---|---|---|
| **30** | Surviving sepsis campaign international guidelines for the management of septic shock and sepsis-associated organ dysfunction in children | | | [46] |
| **31** | Updated nomenclature of delirium and acute encephalopathy: statement of ten Societies | | | [47] |
| **32** | The cardiac arrest centre for the treatment of sudden cardiac arrest due to presumed cardiac cause – aims, function and structure: Position paper of the Association for Acute CardioVascular Care of the European Society of Cardiology (AVCV), European Association of Percutaneous Coronary Interventions (EAPCI), European Heart Rhythm Association (EHRA), European Resuscitation Council (ERC), European Society for Emergency Medicine (EUSEM) and European Society of Intensive Care Medicine (ESICM) | | | [48] |
| **33** | Fluid therapy in neurointensive care patients: ESICM consensus and clinical practice recommendations | | | [49] |
| **34** | Guidelines for the diagnosis and management of critical illness-related corticosteroid insufficiency (CIRCI) in critically ill patients (Part II): Society of Critical Care Medicine (SCCM) and European Society of Intensive Care Medicine (ESICM) 2017 | | 2017 | [50] |
| **35** | Second consensus on the assessment of sublingual microcirculation in critically ill patients: results from a task force of the European Society of Intensive Care Medicine | | | [51] |
| **36** | Guidelines for the Diagnosis and Management of Critical Illness-Related Corticosteroid Insufficiency (CIRCI) in Critically Ill Patients (Part I): Society of Critical Care Medicine (SCCM) and European Society of Intensive Care Medicine (ESICM) 2017 | | 2017 | [52] |
| **37** | International ERS/ESICM/ESCMID/ALAT guidelines for the management of hospital-acquired pneumonia and ventilator-associated pneumonia | | | [53] |
| **38** | An Official American Thoracic Society/European Society of Intensive Care Medicine/Society of Critical Care Medicine Clinical Practice Guideline: Mechanical Ventilation in Adult Patients with Acute Respiratory Distress Syndrome | | | [54] |
| **39** | Early enteral nutrition in critically ill patients: ESICM clinical practice guidelines | | | [55] |
| **40** | Management of severe perioperative bleeding - Guidelines from the European Society of Anaesthesiology | | | [56] |
| **41** | European Society of Anaesthesiology and European Board of Anaesthesiology guidelines for procedural sedation and analgesia in adults | | | [57] |

***Supplementary Table 1: Guidelines Included in Relevance Screening.*** *"AWMF Register No" indicates the entry number of the guideline in the German guideline register of the Arbeitsgemeinschaft der Wissenschaftlichen Medizinischen Fachgesellschaften (AWMF). Version indicates the version number and/or date, if applicable.*

#### 1.3 Encoding Recommendations Using FHIR

We followed a structured process for encoding the recommendations in FHIR based on our previous work [58]. First, an experienced intensive care physician and a medical computer scientist jointly separated the recommendations into a textual representation of the population and the intervention, respectively, and then identified all individual medical concepts that were required to exhaustively characterize the population and intervention. These were then mapped to concepts from the *Systematized Nomenclature of Medicine - Clinical Terms* (SNOMED CT) [59], *Logical Observation Identifiers Names and Codes* (LOINC) [60] and *International Classification of Diseases 10$^{th}$ revision* (ICD-10) [61] vocabularies. We then performed a review session together with additional intensive care physicians and medical computer scientists, during which we discussed the extraction and mappings and performed changes until consensus was reached [58]. Then, a medical information specialist implemented the documented recommendation in Clinical Practice Guidelines (CPG)-on-Evidence-based Medicine (EBM)onFHIR, a FHIR-based format for the computer-interpretable representation of clinical practice guidelines [62], using FHIR ShortHand (FSH) [63]. After syntactic validation via successful conversion with FSH Sushi to JSON and error-free output of the FHIR Java Validator [64], we performed a joint review session among medical computer scientists together with a board-certified intensive care physician, where we validated that the documented specification of the recommendation was faithfully transferred to FSH. Additional validation and alignment of the documented specification of the recommendation was conducted with three independent board-certified intensive care physicians.

The converted recommendations contained a total of 31 different medical concepts (four demographics, eight conditions, three laboratory values, five ventilation observables, eight drugs, two procedures and one episode of care type; **Supplementary Table 2**).

#### 1.4 Execution of FHIR-encoded Guidelines

After retrieval of the required FHIR resources from a separate FHIR server, the execution engine parses the resources and constructs an internal representation of the recommendations in form of a directed acyclic graph, where the upstream nodes represent individual, independent criteria, such as the occurrence of a disease or a certain laboratory value (**Supplementary Figure 1**). The execution engine generates SQL statements that retrieve all time intervals during which the criterion is fulfilled or not fulfilled for each patient from the OMOP CDM database. These intervals are then passed to downstream nodes, which perform either a modification of data from a single upstream node, such as inversion (for exclusion criteria), or integration of data from multiple upstream nodes, such as intersecting the intervals of criteria that are required by the recommendation to be simultaneously present. Ultimately, the criteria that together define the population of the recommendation are intersected with the criteria that define the intervention to yield a complete timeline of intervals during which the recommendation was adhered to or not.

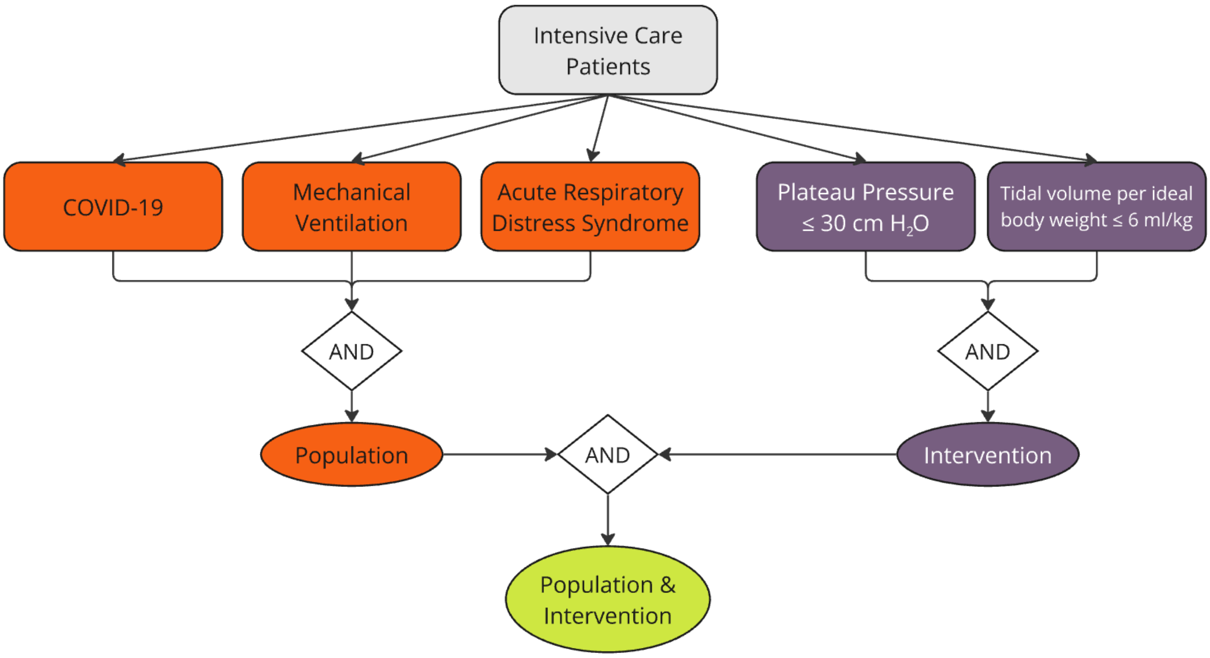


***Supplementary Figure 1 Internal Representation of a Clinical Recommendation in the Execution Engine.*** *Shown is the directed acyclic graph used by the execution engine to represent a clinical recommendation for recommendation #35 ("Tidal Volume"). Each node represents a criterion (rounded rectangle) or logical operation (diamond) required for the recommendation, with colored nodes indicating the type of criteria or category. The grey node represents the initial set of patients that are considered during subsequent steps (here: all patients that were on any intensive care unit during a specific period). Orange nodes represent those criteria that describe which patients are addressed by the recommendation, i.e. the population. Purple nodes represent the criteria that describe what should be performed with the patients, i.e. the intervention criteria (e.g., respiratory system parameters). Logical operators (white) combine these criteria to determine whether the recommendation applies to a patient and whether it is fulfilled. The ellipses at the bottom indicate the final decision points for applicability (population) and adherence (intervention).*

#### 1.5 User Interface for Quality Management

Based on our previous work on a user interface for guideline adherence monitoring [65], we conducted a consensus workshop with eight intensive care professionals with diverse experience levels from assistant to deputy department chair, two medical computer scientists and an expert for human factors engineering in the medical field. The goal for the workshop was to develop a generic user interface that is optimized to monitor guideline adherence in general, not only focused on the recommendations selected in this project.

Therefore, we individually assessed all top 20 ranked recommendations from the recommendation selection step and discussed whether the previously developed user interface allows to efficiently visualize the adherence to the respective recommendation until we reached consensus towards the decisions "allows visualization", "allows visualization after modifications", or "does not allow visualization". In the two latter cases, we collected the changes that were necessary in the view of the participants to effectively visualize the respective recommendation. We collected and prioritized a list of changes and implemented these in our user interface. We then, in an iterative approach, distributed a demo of the user interface to ten intensive care physicians including those that participated in the workshop and collected and integrated their feedback, until no further changes were requested from this focus group.

The final user interface, developed in RShiny [66], visualizes the results generated by the execution engine and supports supervision and quality management workflows (**Supplementary Figure 2**). It provides an overview of guideline adherence for individual patients as well as aggregated data at the ward level. The interface allows users to review the patient-specific data used in each decision, providing transparency in adherence assessments. This functionality was designed to support quality management and improve clinical decision-making in intensive care settings.

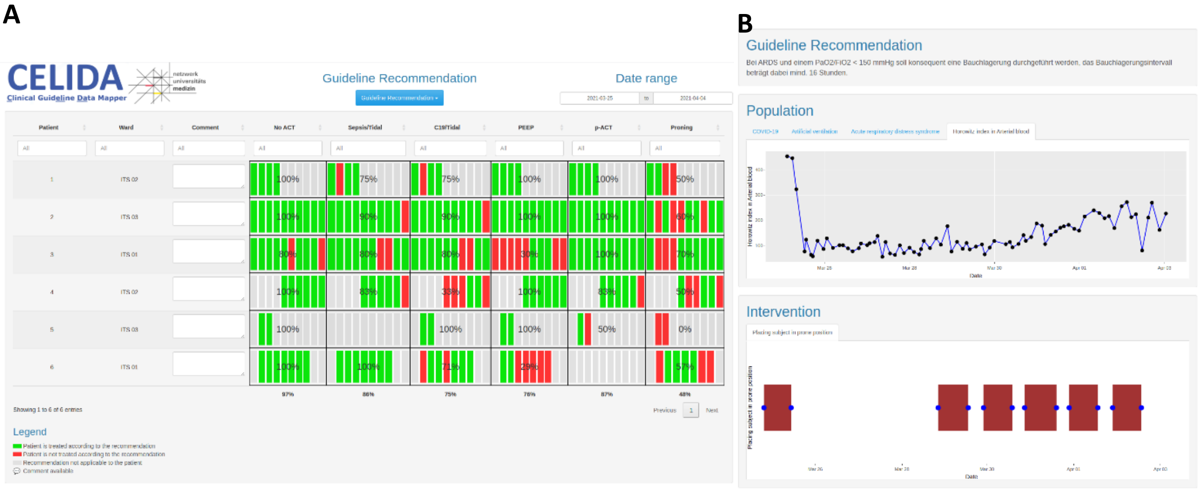


***Supplementary Figure 2 User Interface for Monitoring Guideline Adherence.*** *This figure illustrates the user interface developed for monitoring individual patient treatment adherence to clinical practice guidelines.* ***A*** *The interface displays a dashboard that summarizes the adherence status for all patients on a ward, indicating whether each recommendation is applicable and whether it has been fulfilled. Each row represents one patient on the ward, and each column corresponds to a different recommendation. Within each cell, grey bars represent days when the recommendation is not applicable to the patient, while red and green bars indicate days when the recommendation is applicable but not adhered to (red) or adhered to (green).* ***B*** *Additional features include the ability to drill down into patient-specific details, showing the raw clinical data used by the system to evaluate each recommendation.*

#### 1.6 Information System Vendors in the Participating Hospitals

- Charité – Universitätsmedizin Berlin: COPRA6 (COPRA System GmbH, Germany), i.s.h.med (SAP Deutschland SE & Co. KG, Germany)
- Universitätsmedizin Greifswald: Dräger ICM (Drägerwerk AG & Co. KGaA, Germany)
- Klinikum rechts der Isar / Technische Universität München: QCare ICU (Health Information Management GmbH, Germany), i.s.h.med (SAP Deutschland SE & Co. KG, Germany)
- Universitätsklinikum Würzburg: COPRA6 (COPRA System GmbH, Germany), i.s.h.med (SAP Deutschland SE & Co. KG, Germany)
- Universitätsklinikum Augsburg: ORBIS (Dedalus S.p.A., Italy)

#### 1.7 Validation Study – Selection of Review Dates

We selected ten dates for manual review according to the following algorithm with the intention to minimize statistical bias: First, we ran the execution engine for the period from January 2020 to May 2023 and determined for each date in that period the total number of patients, which were treated according to any of the implemented recommendations. This list, ordered descendant by the number of patients, was the candidate list for date selection. We selected the date with the highest number of patients and removed that date from the candidate list as well as all dates that occurred within 60 days before or after that date. Then, the top entry from the remaining candidate list was added to our selection and this was repeated until we had five entries representing five days on our selection list. To additionally include five randomly selected days, we shuffled the candidate list and added five more dates according to the same algorithm, i.e. always removing all dates within 60 days before or after the included date.

#### 1.8 Ethics Approval

This study was approved by the local ethics committees of the participating hospitals (Ethikkommission Universitätsmedizin Greifswald, Chairperson Prof. S. Ulbricht, application number BB 064/23, approval date: November 6, 2023; Ethikausschuss am Campus Virchow-Klinikum, Charité Universitätsmedizin Berlin, Chairperson Dr. E Kaschina, application number EA2/139/20 approval date: July 30, 2020; Ethikkommission Technische Universität München, Chairperson Prof. G. Schmidt, application number 2023-5-S-KH, approval date: January 10, 2023, amendment date: September 9, 2023; Ethikkommission der Universität Würzburg, Chairperson Prof. R. Jahns, application number 20230208 01, approval date: February 20, 2023; Ethikkommission bei der LMU München, Chairperson Prof. R. M. Huber, application number 23-0592, approval date: August 18, 2023).

| Category | Name | Source Concept | | | OMOP Standardized Vocabulary | | | | |
|---|---|---|---|---|---|---|---|---|---|
| | | System | Code | Display | ID | Domain ID | Name | Unit concept ID | Unit |
| Demographics | **Ideal body weight** | LOINC | 50064-5 | Ideal body weight | 3032445 | Measurement | Ideal body weight | 9529 | kg |
| | **Gender** | LOINC | 46098-0 | Sex | - | person. gender_concept_id | - | - | - |
| | **Height** | LOINC | 8302-2 | Body height | 3036277 | Measurement | Body height | 8582 | cm |
| | **Weight** | LOINC | 29463-7 | Body weight | 4099154 | Measurement | Body weight | 9529 | kg |
| Conditions | **COVID-19** | SNOMED CT | 840539006 | Disease caused by Severe acute respiratory syndrome coronavirus 2 | 37311061 | Condition | COVID-19 | | |
| | **Venous Thrombosis** | SNOMED CT | 111293003 | Venous Thrombosis | 444247 | Condition | Venous Thrombosis | | |
| | **HIT 2** | SNOMED CT | 111588002 | Heparin-induced thrombocytopenia with thrombosis | 4009307 | Condition | Heparin-induced thrombocytopenia with thrombosis | | |
| | **Heparin Allergy** | SNOMED CT | 294872001 | Allergy to heparin | 4169185 | Observation | Allergy to heparin | | |
| | **Heparinoid Allergy** | SNOMED CT | 294876003 | Allergy to heparinoid | 4170358 | Observation | Allergy to heparinoid | | |
| | **Pulmonary Embolism** | SNOMED CT | 59282003 | Pulmonary embolism | 440417 | Condition | Pulmonary embolism | | |
| | **ARDS** | SNOMED CT | 67782005 | Acute respiratory distress syndrome | 4195694 | Condition | Acute respiratory distress syndrome | | |
| | **Atrial Fibrillation** | SNOMED CT | 49436004 | Atrial fibrillation | 313217 | Condition | Atrial fibrillation | | |
| Laboratory | **D-Dimer** | LOINC | 48066-5 | Fibrin D-dimer DDU [Mass/volume] in Platelet poor plasma | 3048530 | Measurement | Fibrin D-dimer DDU [Mass/volume] in Platelet poor plasma | 8842 | ng/mL |
| | **aPTT** | LOINC | 3173-2 | aPTT in Blood by Coagulation assay | 3013466 | Measurement | aPTT in Blood by Coagulation assay | 8555 | s |
| | **Oxygenation Index** | LOINC | 50984-4 | Horowitz index in Arterial blood | 3029943 | Measurement | Horowitz index in Arterial blood | 8876 | mm[Hg] |
| Ventilation Observable | **FiO2** | LOINC | 3150-0 | Inhaled oxygen concentration | 3020716 | Measurement | Inhaled oxygen concentration | 8554 | % |
| | **PaO2** | LOINC | 2703-7 | Oxygen [Partial pressure] in Arterial blood | 3027801 | Measurement | Oxygen [Partial pressure] in Arterial blood | 8876 | mm[Hg] |
| Drugs | **Heparin** | ATC DE | B01AB01 | Heparin | 1367571 | Drug | heparin | 8510 | IE |
| | **Argatroban** | ATC DE | B01AE03 | Argatroban | 1322207 | Drug | argatroban | 8576 | mg |
| | **Dalteparin** | ATC DE | B01AB04 | Dalteparin | 1301065 | Drug | dalteparin | 8510 | IE |
| | **Enoxaparin** | ATC DE | B01AB05 | Enoxaparin | 1301025 | Drug | enoxaparin | 8576 | mg |
| | **Nadroparin** | ATC DE | B01AB06 | Nadroparin | 19001014 | Drug | nadroparin | 8510 | IE |
| | **Certoparin** | ATC DE | B01AB13 | Certoparin | 19016072 | Drug | certoparin | 8510 | IE |

| | | | | | | | | | |
|---|---|---|---|---|---|---|---|---|---|
| | **Fondaparinux** | ATC DE | B01AX05 | Fondaparinux | 1315865 | Drug | fondaparinux | 8576 | mg |
| | **Tinzaparin** | ATC DE | B01AB10 | Tinzaparin | 1308473 | Drug | tinzaparin | 8510 | IE |
| **Procedure** | **Ventilation** | SNOMED CT | 40617009 | Artificial respiration | 4230167 | Procedure | Artificial respiration | | |
| | **Prone Positioning** | SNOMED CT | 431182000 | Placing subject in prone position | 4196006 | Procedure | Placing subject in prone position | | |
| **Ventilator Management** | **Tidal Volume** | LOINC | 76222-9 | Tidal volume^on ventilator | 21490854 | Measurement | Tidal volume Ventilator --on ventilator | 8587 | ml |
| | **End-inspiratory airway pressure** | LOINC | 76259-1 | Pressure.plateau Respiratory system airway --on ventilator | 36303946 | Measurement | Pressure.plateau Respiratory system airway --on ventilator | 44777590 | cm[H2O] |
| | **PEEP** | LOINC | 76248-4 | PEEP Respiratory system --on ventilator | 21490855 | Measurement | PEEP Respiratory system --on ventilator | 44777590 | cm[H2O] |
| **Episode of Care** | **Intensive Care** | kontaktart-de | intensivstationaer | Intensivstationär | 32037 | Visit | Intensive Care | | |

***Supplementary Table 2: Clinical Features Used for Modeling Recommendations.*** *Shown are the clinical features used to model the selected recommendations in the execution engine. The features are categorized based on their clinical category (e.g., demographics, conditions). For each feature, the corresponding source concept is shown, including its code system (e.g., LOINC, SNOMED CT), code, and display name. Also shown is the mapping of each concept to the OMOP standardized vocabulary, including the OMOP domain, concept ID, and, where applicable, unit of measurement.*

| Concept | Hospital a | | | Hospital b | | | Hospital c | | | Hospital d | | | Hospital e | | |
|---|---|---|---|---|---|---|---|---|---|---|---|---|---|---|---|
| | Unique Patients | n (avg) | Average Duration | Unique Patients | n (avg) | Average Duration | Unique Patients | n (avg) | Average Duration | Unique Patients | n (avg) | Average Duration | Unique Patients | n (avg) | Average Duration |
| Acute respiratory distress syndrome | 58 | 1.0 | 10.3 days | 500 | 1.5 | 28.1 days | 356 | 2.7 | 15.8 days | 170 | 1.0 | 38.0 days | 839 | 1 | 22.3 days |
| Mechanical Ventilation | 1894 | 3.2 | 1.6 days | 21471 | 25.3 | 4.0 days | 5027 | 1.2 | 3.1 days | 3543 | 5.1 | 1.8 days | 9988 | 9.9 | 0.7 days |
| Atrial fibrillation | 1053 | 1.0 | 5.7 days | 2971 | 1.2 | 7.5 days | 919 | 2.2 | 21.4 days | 1384 | 1.1 | 28.9 days | 902 | 1.2 | 14.8 days |
| COVID-19 | 283 | 1.3 | 9.0 days | 627 | 1.4 | 12.8 days | 658 | 3.5 | 15.1 days | 471 | 1.0 | 30.7 days | 1188 | 1 | 17.6 days |
| Intensive Care | 5067 | 1.0 | 5.0 days | 18372 | 1.7 | 4.6 days | 8642 | 1.3 | 3.6 days | 6614 | 1.9 | 5.5 days | 18812 | 1.2 | 3.3 days |
| Pulmonary embolism | 87 | 1.0 | 5.3 days | 689 | 1.3 | 10.9 days | 79 | 2.7 | 23.3 days | 356 | 1.0 | 33.3 days | 466 | 1.1 | 19.8 days |
| Venous thrombosis | 130 | 1.1 | 7.6 days | 636 | 1.2 | 14.9 days | 51 | 2.4 | 23.0 days | 269 | 1.1 | 41.9 days | 552 | 1.2 | 30.8 days |

***Supplementary Table 3: Occurrences of conditions, procedures and visits for each hospital**. Shown are the number of unique patients for whom the respective condition was recorded (Unique Patients), the average number of times the condition appeared per patient for those patients for whom the condition was recorded (n (avg)), as well as the average duration of each concept in the datasets.*

| Concept (unit) | Clinic a | | | Clinic b | | | Clinic c | | | Clinic d | | | Clinic e | | |
|---|---|---|---|---|---|---|---|---|---|---|---|---|---|---|---|
| | Unique Patients | n (avg) | median [IQR] | Unique Patients | n (avg) | median [IQR] | Unique Patients | n (avg) | median [IQR] | Unique Patients | n (avg) | median [IQR] | Unique Patients | n (avg) | median [IQR] |
| **Body height (cm)** | 4073 | 1.0 | 171 [165-180] | 18853 | 1.3 | 170 [160-178] | 205 | 1.1 | 175 [166-180] | 5622 | 7.8 | 172 [165-180] | 15659 | 1.0 | 172 [165-178] |
| **Body weight (kg)** | 4099 | 1.0 | 80 [70-90] | 21439 | 4.2 | 55.0 [18.8-77.0] | 3225 | 2.2 | 71 [65-80] | 6350 | 87.0 | 80 [70-90] | 16090 | 1.0 | 80 [68-90] |
| **Fibrin D-dimer DDU [Mass/volume] in Platelet poor plasma (ng/mL)** | 597 | 4.4 | 3.9 [1.8-8.6] | 3005 | 9.1 | 2.4 [0.9-7.3] | 777 | 3.4 | 3.0 [1.4-6.6] | 120 | 2.2 | 4068 [1314-15210] | 2565 | 4.0 | 4.2 [1.8-10.1] |
| **Horowitz index in Arterial blood (mm[Hg])** | 1827 | 40.1 | 254 [174-334] | 16309 | 44.2 | 297.2 [219.7-384.4] | 2699 | 13.1 | 196.9 [119.3-282.5] | 3165 | 116.0 | 238 [174-305] | 9425 | 30.2 | 214 [153-284] |
| **Ideal body weight (kg)** | 4073 | 1.0 | 52.3 [43.2-61.4] | 18853 | 1.3 | 63.3 [53.3-73.3] | 204 | 1.1 | 70.6 [62.8-75.1] | 5520 | 7.8 | 64 [56-73] | 15659 | 1.0 | 66.0 [57.0-73.3] |
| **Inhaled oxygen concentration (%)** | 1993 | 1015.6 | 35 [25-50] | 21544 | 228.2 | 40 [31-52] | 3775 | 27.4 | 41 [39-60] | 5827 | 666.5 | 35 [29-41] | 9513 | 197.1 | 40.0 [34.8-50.0] |
| **Oxygen [Partial pressure] in Arterial blood (mm[Hg])** | 4359 | 60.0 | 84.0 [72.9-98.3] | 19412 | 60.8 | 97.1 [80.6-120.0] | 6137 | 14.2 | 85.3 [68.1-107.0] | 5357 | 117.9 | 88.3 [76.0-103.1] | | | |
| **PEEP Respiratory system --on ventilator (cm[H2O])** | 1927 | 244.1 | 7.4 [6.2-8.5] | 19235 | 117.1 | 7 [5-8] | 3771 | 27.2 | 7.0 [5.0-9.6] | 3479 | 578.1 | 6.3 [5.2-8.2] | 9542 | 191.1 | 7.9 [5.3-10.1] |
| **Pressure.plateau Respiratory system airway --on ventilator (cm[H2O])** | 1595 | 165.7 | 19.4 [16.3-22.4] | 19636 | 178.6 | 19 [16-24] | 3734 | 24.9 | 17 [13-22] | 3479 | 578.2 | 17.1 [12.2-22.6] | 9854 | 190.0 | 20.9 [16.8-25.9] |
| **Tidal volume Ventilator --on ventilator (mL)** | 1957 | 248.9 | 459 [383-543] | 17574 | 197.7 | 450 [356-542] | 3732 | 24.7 | 479 [399-568] | 3117 | 624.3 | 436.8 [364.4-520.2] | 9767 | 190.3 | 480 [405-573] |
| **aPTT in Blood by Coagulation assay (s)** | 4911 | 6.7 | 25 [22-29] | 23402 | 17.2 | 37.8 [32.0-47.2] | 5750 | 4.8 | 37 [31-47] | 6562 | 61.9 | 33 [29-41] | 18687 | 8.0 | 28.2 [23.6-37.9] |

***Supplementary Table 4: Measurement statistics for each hospital.*** *Shown are the number of unique patients for which measurements of the respective type were recorded (Unique Patients), the mean number of valid measurements per patient of those patients for who any measurement was recorded (n (avg)) as well as the median and IQR of the values.*

| | | Clinic a | | | Clinic b | | | Clinic c | | | Clinic d | | | Clinic e | | |
|---|---|---|---|---|---|---|---|---|---|---|---|---|---|---|---|---|
| Drug (Unit) | Route | Unique Patients | n (Avg) | median [IQR] | Unique Patients | n (Avg) | median [IQR] | Unique Patients | n (Avg) | median [IQR] | Unique Patients | n (Avg) | median [IQR] | Unique Patients | n (Avg) | median [IQR] |
| **apixaban (mg)** | **p.o.** | 42 | 10.6 | 2.5 [2.5-5.0] | | | | | | | | | | | | |
| **argatroban (mg)** | **i.v.** | 1 | 2.0 | 2.8 [2.5-3.1] | 423 | 67.1 | 14.5 [5.4-32.3] | 61 | 67.3 | 5.5 [1.9-12.9] | 62 | 118.8 | 0 [0-24] | 89 | 27.6 | 22.9 [7.8-50.0] |
| | **s.c.** | | | | 29 | 2.6 | 5 [1-20] | | | | | | | 8 | 1.2 | 3.0 [2.1-5.0] |
| **certoparin ([U])** | **i.v.** | | | | | | | | | | 42 | 4.4 | 3000 [3000-3000] | | | |
| | **s.c.** | 1056 | 6.0 | 3000 [3000-3000] | | | | | | | 1548 | 6.8 | 3000 [3000-3000] | 2 | 1.0 | 8000 [8000-8000] |
| **dabigatran (mg)** | **p.o.** | 2 | 12.0 | 110 [110-110] | | | | | | | | | | | | |
| **dalteparin ([U])** | **s.c.** | 2319 | 7.2 | 5000 [5000-5000] | 1662 | 7.0 | 5000 [5000-7500] | 73 | 7.2 | 5000 [5000-5000] | | | | | | |
| **enoxaparin (mg)** | **i.v.** | | | | | | | 182 | 60.7 | 4.0 [2.2-6.1] | 14 | 3.1 | 40 [20-40] | 1 | 1.0 | 60 [60-60] |
| | **s.c.** | 118 | 7.2 | 60 [40-80] | 404 | 16.7 | 40 [40-60] | 8 | 7.2 | 40 [40-40] | 2942 | 12.9 | 40 [40-40] | 8627 | 5.5 | 40 [40-60] |
| **fondaparinux (mg)** | **i.v.** | | | | | | | | | | 2 | 2.5 | 2.5 [2.5-2.5] | | | |
| | **s.c.** | | | | 80 | 5.8 | 2.5 [2.5-5.0] | | | | 53 | 5.8 | 2.5 [2.5-2.5] | 618 | 2.5 | 2.5 [2.5-2.5] |
| **heparin ([U])** | **i.v.** | 376 | 9.9 | 4000 [1667-8233] | 7760 | 24.0 | 4583 [1875-8871] | 2313 | 50.0 | 1440 [707-2652] | 963 | 76.1 | 0 [0-5600] | 7192 | 15.2 | 77378 [38333-133704] |
| | **s.c.** | 17 | 3.6 | 5000 [5000-5000] | 2411 | 2.0 | 5000 [2000-5000] | 4 | 44.2 | 5000 [1-5000] | 2 | 4.5 | 0 [0-0] | 1618 | 3.3 | 0 [0-5000] |
| **nadroparin ([U])** | **s.c.** | | | | 120 | 4.3 | 3800 [2850-5700] | | | | | | | 1 | 5.0 | 2850 [2850-2850] |
| **tinzaparin ([U])** | **s.c.** | 206 | 6.7 | 14000 [13000-16000] | 7 | 4.9 | 10000 [10000-10000] | 11 | 6.1 | 14000 [9000-20000] | | | | 4 | 7.5 | 14000 [14000-14000] |

***Supplementary Table 5: Drug statistics for each hospital**. Shown are the number of unique patients that received the respective drug (Unique Patients), the average number of drug administrations per patient of those patients that received the respective drug (n (Avg)) as well as the median and IQR of the applied doses.*

| | Clinic a | | Clinic b | | Clinic c | |
|---|---|---|---|---|---|---|
| | **Population** | **Intervention** | **Population** | **Intervention** | **Population** | **Intervention** |
| Decisions Performed by Multiple Physicians | 336 | 88 | 924 | 112 | 534 | 143 |
| Rating Physicians | 2 | 2 | 3 | 3 | 2 | 2 |
| Percent Agreement (%) | 80,7% | 58,4% | 95,5% | 56,7% | 83.5% | 82.8% |

***Supplementary Table 6: Inter-Rater Reliability*** *among physician reviewers involved in the validation of system-generated decisions. The number of cases that were reviewed by multiple rating physicians is reported, along with the total number of physicians who participated in the evaluation. Percent agreement indicates the level of concordance between physicians, providing an indication of the consistency and reliability of the manual reviews across different sites.*

### 15 - Prophylactic Anticoagulation (English)

#### Recommendation

##### Guideline

S3 guideline Recommendations for the inpatient treatment of patients with COVID-19 – Living Guideline - https://register.awmf.org/de/leitlinien/detail/113-001LG

##### Summary

###### English (machine translation)

Hospitalized patients with COVID-19 should receive standard drug thromboembolism prophylaxis with low molecular weight heparin in the absence of contraindications. Alternatively, fondaparinux may be used.

##### Justification

###### English (machine translation)

Thromboembolic events are a common complication of COVID-19 and predominantly affect the venous but also the arterial vasculature (112-114). Therefore, all hospitalized patients should receive low-molecular-weight heparin (NMH) for prophylaxis of venous thromboembolism (VTE) at a dosage approved for high-risk settings, e.g., enoxaparin 4,000 IU, dalteparin 5,000 IU, tinzaparin 4,500 IU, certoparin 3,000 IU, nadroparin (< 70kg bw 3,800 IU, > 70kg bw 5,700 IU). Alternatively, e.g. in cases of heparin intolerance or previous heparin-induced thrombocytopenia (HIT) administration of fondaparinux is possible. The recommendation is based on the S3 guideline Prophylaxis of venous thromboembolism (115). The benefit of drug-based VTE prophylaxis, preferably with NMH, in hospitalized patients with acute internal diseases and bed confinement, has been prospectively investigated in several randomized trials. Regarding hospitalized patients with COVID-19, there are no specific trial data on VTE prophylaxis; however, the evidence available to date is applicable to the pandemic situation. Considering contraindications, thromboprophylaxis with NMH (or alternatively with fondaparinux) is highly effective in reducing the risk of VTE without significantly increasing the risk of major bleeding.

> Note: Drug doses for determining whether an anticoagulant dose is therapeutic can be found on 18 - No Therapeutic Anticoagulation (English)

#### Population

| | | Inclusion ⊕ / Exclusion ⊖ | Name | Category | definition.type | definition.value |
|---|---|---|---|---|---|---|
| PopHospitalisedCOVID19PatientsWOVenousThrombosisWOCI | Population for recommendation 15: Hospitalised COVID-19 patients without (venous) thrombosis and without contraindications to LWMH. | ⊕ | COVID-19 | Condition | SCT 404684003 "Clinical finding (finding)" | $sct#840539006 "Disease caused by Severe acute respiratory syndrome coronavirus 2 (disorder)" |
| | | ⊖ *any-of* | HIT 2 | Condition | SCT 404684003 "Clinical finding (finding)" | $sct#111588002 "Heparin-induced thrombocytopenia with thrombosis (disorder)" |
| | | | Heparin Allergy | Allergy | SCT 609328004 "Allergic disposition (finding)" | $sct#294872001 "Allergy to heparin (finding)" |
| | | | Heparinoid Allergy | Allergy | SCT 609328004 "Allergic disposition (finding)" | $sct#294876003 "Allergy to heparinoid (finding)" |
| PopHospitalisedCOVID19PatientsWOVenousThrombosisWITHCI | Population for recommendation 15: Hospitalised COVID-19 patients without (venous) thrombosis, existing contraindications to LWMH. | ⊕ | COVID-19 | Condition | SCT 404684003 "Clinical finding (finding)" | $sct#840539006 "Disease caused by Severe acute respiratory syndrome coronavirus 2 (disorder)" |

| | | | | | | |
|---|---|---|---|---|---|---|
| | | *any-of* | HIT 2 | Condition | SCT 404684003 "Clinical finding (finding)" | $sct#111588002 "Heparin-induced thrombocytopenia with thrombosis (disorder)" |
| | | | Heparin Allergy | Allergy | SCT 609328004 "Allergic disposition (finding)" | $sct#294872001 "Allergy to heparin (finding)" |
| | | | Heparinoid Allergy | Allergy | SCT 609328004 "Allergic disposition (finding)" | $sct#294876003 "Allergy to heparinoid (finding)" |

### Intervention

Note: Drug doses for determining whether an anticoagulant dose is therapeutic can be found on 18 - No Therapeutic Anticoagulation (English)

| Name | Description | Population | | | Actions/Activities | | | | | |
|---|---|---|---|---|---|---|---|---|---|---|
| | | | selection-behaviour | selection-behaviour | Name | Action Category | productCodeableConcept | Route | Drug Dosage | perform doNotP |
| AntithrombLWMHProphIn-HospitalisedCOVID19PatientsRecommPlan | Antithrombotic prophylaxis with LWMH in hospitalised COVID-19 patients | PopHospitalised<br>COVID19-PatientsWOVenousThrombosisWOCI | #all | #exactly (1) | ProphylacticAnticoagulationWDalteparin | drugAdministration | $atcde#B01AB04 "Dalteparin"<br>$sct#372563008 "Dalteparin (substance)" | subcutaneous | • 2500 IU/d<br>• 5000 IU/d | |
| | | | | | ProphylacticAnticoagulationWEnoxaparin | drugAdministration | $atcde#B01AB05 "Enoxaparin"<br>$sct#372562003 "Enoxaparin (substance)" | subcutaneous | • 20 mg/d<br>• 40 mg/d | |
| | | | | | ProphylacticAnticoagulationWNadroparinLowWeight | drugAdministration | $atcde#B01AB06 "Nadroparin"<br>$sct#699946002 "Nadroparin (substance)" | subcutaneous | Weight (kg) / Dose (1x/d)<br>70kg: 3800 IE<br>>70kg: 5700 IE | |
| | | | | | ProphylacticAnticoagulationWCertoparin | drugAdministration | $atcde#B01AB13 "Certoparin"<br>$sct#395961003 "Certoparin (substance)" | subcutaneous | = 3000 IE/d | |
| | | | | | ProphylacticAnticoagulationWTinzaparin | drugAdministration | $atcde#B01AB10 "Tinzaparin"<br>$sct#412608008 "Tinzaparin (substance)" | subcutaneous | = 3.500 IE | |
| | | | | | ProphylacticAnticoagulationWHeparinSC | drugAdministration | $atcde#B01AB01 "Heparin"<br>$sct#372877000 "Heparin (substance)" | subcutaneous | • 5000 IE 2x/day<br>• 5000 IE 3x/day<br>• 7500 IE 2x/day | |
| | | | | #all | NoTherapeuticAnticoagulationWDalteparin1xd | drugAdministration | $atcde#B01AB04 "Dalteparin"<br>$sct#372563008 "Dalteparin (substance)" | subcutaneous | =200 IE/kg 1x/d | |
| | | | | | NoTherapeuticAnticoagulationWDalteparin2xd | drugAdministration | $atcde#B01AB04 "Dalteparin"<br>$sct#372563008 "Dalteparin (substance)" | subcutaneous | =100 IE/kg 2x/d | |

<table>
<tr><td></td><td></td><td></td><td></td><td></td><td>TherapeuticAnticoagulationNMHEnoxaparin2xd</td><td>drugAdministration</td><td>$atcde#B01AB05 "Enoxaparin"<br><br>$sct#372562003 "Enoxaparin (substance)"</td><td>subcutaneous</td><td><table><tr><th>Weight (kg)</th><th>Daily Dose (IE)</th></tr><tr><td>46 - 56</td><td>10.000</td></tr><tr><td>57 - 68</td><td>12.500</td></tr><tr><td>69 - 82</td><td>15.000</td></tr><tr><td>> 83</td><td>18.000</td></tr></table></td><td></td></tr>
<tr><td></td><td></td><td></td><td></td><td></td><td>NoTherapeuticAnticoagulationWEnoxaparin1xd</td><td>drugAdministration</td><td>$atcde#B01AB05 "Enoxaparin"<br><br>$sct#372562003 "Enoxaparin (substance)"</td><td>subcutaneous</td><td>1.5 mg/kg; 1x/d</td><td></td></tr>
<tr><td></td><td></td><td></td><td></td><td></td><td>TherapeuticAnticoagulationNMHEnoxaparin2xd</td><td>drugAdministration</td><td>$atcde#B01AB05 "Enoxaparin"<br><br>$sct#372562003 "Enoxaparin (substance)"</td><td>subcutaneous</td><td>1 mg/kg ; 2x/d</td><td></td></tr>
<tr><td></td><td></td><td></td><td></td><td></td><td>NoTherapeuticAnticoagulationWNadroparin</td><td>drugAdministration</td><td>$atcde#B01AB06 "Nadroparin"<br><br>$sct#699946002 "Nadroparin (substance)"</td><td>subcutaneous</td><td><ul><li>2x/d according to:</li></ul><table><tr><th>Weight (kg)</th><th>Dose s.c. (IE; 2x/d)</th></tr><tr><td>< 50</td><td>3800</td></tr><tr><td>50 bis 59</td><td>4750</td></tr><tr><td>60 bis 69</td><td>5700</td></tr><tr><td>70 bis 79</td><td>6650</td></tr><tr><td>80 bis 89</td><td>7600</td></tr><tr><td>90</td><td>8550</td></tr></table></td><td></td></tr>
<tr><td></td><td></td><td></td><td></td><td></td><td>NoTherapeuticAnticoagulationWCertoparin</td><td>drugAdministration</td><td>$atcde#B01AB13 "Certoparin"<br><br>$sct#395961003 "Certoparin (substance)"</td><td>subcutaneous</td><td>8000 IE 2x/d</td><td></td></tr>
<tr><td></td><td></td><td></td><td></td><td></td><td rowspan="2">NoTherapeuticAnticoagulationWUFHOrArgatra</td><td>drugAdministration</td><td>$atcde#B01AB01 "Heparin"<br><br>$sct#372877000 "Heparin (substance)"</td><td>intravenous</td><td></td><td></td></tr>
<tr><td></td><td></td><td></td><td></td><td></td><td>drugAdministration</td><td>$atcde#B01AE03 "Argatroban"<br><br>$sct#116508003 "Argatroban (substance)"</td><td>intravenous</td><td></td><td></td></tr>
<tr><td></td><td></td><td></td><td></td><td></td><td>NoTherapeuticAnticoagulationWTinzaparin</td><td>drugAdministration</td><td>$atcde#B01AB10 "Tinzaparin"<br><br>$sct#412608008 "Tinzaparin (substance)"</td><td>subcutaneous</td><td>175 IE/kg 1x/d</td><td></td></tr>
<tr><td></td><td></td><td></td><td></td><td></td><td></td><td></td><td></td><td></td><td></td><td></td></tr>
<tr><td>AntithrombFondapProphIn-HospitalisedCOVID19PatientsRecommPlan</td><td>Antithrombotic prophylaxis with Fondaparinux in hospitalized COVID-19 patients</td><td>PopHospitalised<br><br>COVID19-PatientsWOVenousThrombosisWITHCI</td><td rowspan="2">#alll</td><td></td><td>AntithromboticProphylaxisFondaparinuxSubcutaneous</td><td>drugAdministration</td><td>$atcde#B01AX05 "Fondaparinux"<br><br>$sct#708189008 "Fondaparinux (substance)"</td><td></td><td>=2.5 'mg'/day</td><td></td></tr>
<tr><td></td><td></td><td></td><td></td><td>TherapeuticAnticoagulationFondaparinux</td><td>drugAdministration</td><td>$atcde#B01AX05 "Fondaparinux"<br><br>$sct#708189008 "Fondaparinux (substance)"</td><td>subcutaneous</td><td><table><tr><th>Weight (kg)</th><th>Dose (1x/d)</th></tr><tr><td>< 50</td><td>5 mg</td></tr><tr><td>>= 50, <= 100</td><td>7.5 mg</td></tr><tr><td>> 100</td><td>10 mg</td></tr></table></td><td></td></tr>
</table>

### 18 - No Therapeutic Anticoagulation (English)

#### Recommendation

##### Guideline

S3 guideline Recommendations for the inpatient treatment of patients with COVID-19 – Living Guideline - https://register.awmf.org/de/leitlinien/detail/113-001LG

##### Summary

###### English (machine translation)

Therapeutic anticoagulation should not be used in ICU patients without a specific indication (e.g., pulmonary embolism).

##### Justification

###### English (machine translation)

**Presentation of the evidence base**
The evidence base in ICU patients includes four evaluable trials of parenteral anticoagulation (124, 128-130). For the most heavily weighted end point, the occurrence of a thrombotic event or mortality by day 28, the effect estimate based on the REMAP-CAP trial and the HEP-COVID trial (subgroup) has a relative risk of 0.98 (95% CI 0.86-1.12, fixed effect model), indicating that in ICU patients, therapeutic anticoagulation has no effect on the clinically relevant outcome (128, 130). In contrast, the risk of major bleeding is nominally but not significantly increased in the subgroup of COVID-19 patients requiring intensive care (rel. risk 1.85; 95% CI 0.81-4.23) (128, 130). Meta-analysis with studies on all patient collectives showed a significantly increased risk of major bleeding (rel. risk 1.78; 95% CI 1.15-2.74). The ACTION trial (rivaroxaban versus NMH prophylaxis) studied only a small number of patients with severe COVID-19 progression (51 versus 40 patients per group) (126). Overall, however, this study showed similar effects to the REMAP-CAP study of COVID-19 patients requiring intensive care in terms of the occurrence of thrombotic events or mortality by day 28.
For 28-day mortality, three trials in COVID-19 patients requiring intensive care showed a nonsignificant survival benefit with therapeutic anticoagulation (124, 128, 129). The largest study of more than 1000 COVID-19 patients requiring intensive care (130) and a small study of 20 intensive care patients (124) reported hospital mortality as an end point and showed no benefit from therapeutic anticoagulation in meta-analysis for all patients regardless of disease severity.

**The rationale for Level of Recommendation.**
The certainty of the evidence was downgraded for most endpoints because of bias risk and indirectness or because of imprecision or heterogeneity of effect estimates. The largest trial with the highest weight in analyses and with more than 1000 COVID-19 patients requiring intensive care administered anticoagulation at therapeutic doses without explanation in only about 80% of patients in the intervention group (bias risk) and >59% of the comparison group received anticoagulation at semi therapeutic or higher doses (indirectness) (130). Considering the very questionable beneficial effects on clinical outcomes in ICU patients (no significant difference in the occurrence of thromboembolic events or mortality) and signal for increased bleeding, therapeutic anticoagulation should not be given without a specific indication (eg, pulmonary embolism).

##### Reference (proposals by Fridtjof & Gregor) for determining whether an anticoagulant dose is therapeutic

<table>
<tr><th>Drug</th><th>Tradename</th><th>Prophylactic</th><th>Therapeutic</th></tr>
<tr><td>Dalteparin</td><td>Fragmin</td><td>2500 -5000 IE s.c. 1x/d<br><br>(actually: 2500 OR 5000 IE)</td><td><ul><li>100 IE/kg 2x/d</li><li>OR 200 IE/kg 1x/d</li><li>OR table:</li></ul><table><tr><th>Weight (kg)</th><th>Daily Dose (IE)</th></tr><tr><td>46 - 56</td><td>10.000</td></tr><tr><td>57 - 68</td><td>12.500</td></tr><tr><td>69 - 82</td><td>15.000</td></tr><tr><td>> 83</td><td>18.000</td></tr></table></td></tr>
<tr><td>Enoxparin</td><td>Clexane<br><br>Inhixa</td><td>20 - 40 mg/d<br><br>(actually: 20 OR 40 mg/d)</td><td><ul><li>1.5 mg/kg 1x/d</li><li>OR 1 mg/kg 2x/d</li></ul></td></tr>
</table>

<table>
<tr><td>Nadroparin</td><td>Fraxiparin</td><td><table><tr><th>Weight (kg)</th><th>Dose (1x/d)</th></tr><tr><td>70kg</td><td>3800 IE</td></tr><tr><td>>70kg</td><td>5700 IE</td></tr></table></td><td><ul><li>2x/d according to:</li></ul><table><tr><th>Weight (kg)</th><th>Dose s.c. (IE; 2x/d)</th></tr><tr><td>< 50</td><td>3800 IE</td></tr><tr><td>50 bis 59</td><td>4750 IE</td></tr><tr><td>60 bis 69</td><td>5700 IE</td></tr><tr><td>70 bis 79</td><td>6650 IE</td></tr><tr><td>80 bis 89</td><td>7600 IE</td></tr><tr><td>90</td><td>8550 IE</td></tr></table></td></tr>
<tr><td>Certoparin</td><td>Mono-Embolex</td><td>3000 IE/d</td><td>8000 IE 2x/d (alle 12 h)</td></tr>
<tr><td>Tinzaparin</td><td>Innohep</td><td>3500 IE 1x/d</td><td>175 IE/kg 1x/d</td></tr>
<tr><td>Fondaparinux</td><td>Arixtra</td><td>2,5 mg 1x/d</td><td><table><tr><th>Weight (kg)</th><th>Dose (1x/d)</th></tr><tr><td>< 50</td><td>5 mg</td></tr><tr><td>>= 50, <= 100</td><td>7.5 mg</td></tr><tr><td>> 100</td><td>10 mg</td></tr></table></td></tr>
<tr><td>UFH and Argatroban</td><td></td><td>N/A</td><td>apTT > 50 sec.</td></tr>
<tr><td>Heparin subkutan</td><td></td><td><ul><li>5000 IE alle 8-12h (2-3x/d)</li><li>75000 IE all 12h (2x/d)</li></ul></td><td></td></tr>
</table>

**CAVE (=BEWARE) when concerning LWMH:**

level of anti-Xa activity (a blood lab value) should be checked in a blood sample, taken 3-5 hours after the third administration (or after a later administration). This lab value is the actual check for therapeutic effect, and the important test whether the drug is overdosed. Since this therapeutic anticoagulation is hard to model, we decided to model the recommendation as follows: if "more than" "high-level prophylactic anticoagulation" is detected, it is considered therapeutic, and therefore does not fulfill the recommendation.

### Population

<table>
<tr><th>Name</th><th>Description</th><th colspan="5">Criteria</th></tr>
<tr><th></th><th></th><th>Inclusion + / Exclusion −</th><th>Name</th><th>Category</th><th>definition.type</th><th>definition.value</th></tr>
<tr><td rowspan="4">PopHospitalisedICUCOVID19Patients</td><td rowspan="4">Hospitalised COVID-19 patients, treated on ICU, without thrombembolic complication</td><td>+</td><td>COVID-19</td><td>Condition</td><td>SCT 404684003 "Clinical finding (finding)"</td><td>$sct#840539006 "Disease caused by severe acute respiratory syndrome coronavirus 2 (disorder)"</td></tr>
<tr><td>+</td><td>ICU</td><td>episodeOfCare</td><td>LOINC 78030-4 "Episode of care Type"</td><td>kontaktart-de#intensivstationaer</td></tr>
<tr><td>−</td><td>pulmonary embolism</td><td>Condition</td><td>SCT 404684003 "Clinical finding (finding)"</td><td>$sct=#59282003 "Pulmonary embolism (disorder)"</td></tr>
<tr><td>−</td><td>venous thrombosis</td><td>Condition</td><td>SCT 404684003 "Clinical finding (finding)"</td><td>$sct#111293003 "Venous thrombosis (disorder)"</td></tr>
</table>

<table>
<tr><td></td><td></td><td></td><td>atrial fibrillation</td><td>Condition</td><td>SCT 404684003 "Clinical finding (finding)"</td><td>SCT 49436004 "Atrial fibrillation (disorder)"</td></tr>
</table>

### Intervention

Note: Drug doses for determining whether an anticoagulant dose is therapeutic can be found on anticoagulant-doses

<table>
<tr><th>Name</th><th>Population</th><th colspan="7">Actions/Actitivies</th></tr>
<tr><th></th><th></th><th>Name</th><th>Action category</th><th>productCodeableConcept</th><th>Route</th><th>Drug Dosage</th><th>Goal</th><th>perform? (!doNotPerform)</th></tr>
<tr><td rowspan="10">NoTherapeuticAnticoagulationICUCOVID19NoIndicationPlan</td><td rowspan="10">PopHospitalised<br><br>ICUCOVID19Patients</td><td>NoTherapeuticAnticoagulationWDalteparin1xd</td><td>drugAdministration</td><td>$atcde#B01AB04 "Dalteparin"<br><br>$sct#372563008 "Dalteparin (substance)"</td><td>subcutaneous</td><td>=200 IE/kg 1x/d</td><td></td><td></td></tr>
<tr><td>NoTherapeuticAnticoagulationWDalteparin2xd</td><td>drugAdministration</td><td>$atcde#B01AB04 "Dalteparin"<br><br>$sct#372563008 "Dalteparin (substance)"</td><td>subcutaneous</td><td>=100 IE/kg 2x/d</td><td></td><td></td></tr>
<tr><td>TherapeuticAnticoagulationNMHDalteparinTable</td><td>drugAdministration</td><td>$atcde#B01AB04 "Dalteparin"<br><br>$sct#372563008 "Dalteparin (substance)"</td><td>subcutaneous</td><td><table><tr><th>Weight (kg)</th><th>Daily Dose (IE)</th></tr><tr><td>46 - 56</td><td>10.000</td></tr><tr><td>57 - 68</td><td>12.500</td></tr><tr><td>69 - 82</td><td>15.000</td></tr><tr><td>> 83</td><td>18.000</td></tr></table></td><td></td><td></td></tr>
<tr><td>NoTherapeuticAnticoagulationWEnoxaparin1xd</td><td>drugAdministration</td><td>$atcde#B01AB05 "Enoxaparin"<br><br>$sct#372562003 "Enoxaparin (substance)"</td><td>subcutaneous</td><td>1.5 mg/kg; 1x/d</td><td></td><td></td></tr>
<tr><td>TherapeuticAnticoagulationNMHEnoxaparin2xd</td><td>drugAdministration</td><td>$atcde#B01AB05 "Enoxaparin"<br><br>$sct#372562003 "Enoxaparin (substance)"</td><td>subcutaneous</td><td>1 mg/kg ; 2x/d</td><td></td><td></td></tr>
<tr><td>NoTherapeuticAnticoagulationWNadroparin</td><td>drugAdministration</td><td>$atcde#B01AB06 "Nadroparin"<br><br>$sct#699946002 "Nadroparin (substance)"</td><td>subcutaneous</td><td>• 2x/d according to:<table><tr><th>Weight (kg)</th><th>Dose s.c. (IE; 2x/d)</th></tr><tr><td>< 50</td><td>3800</td></tr><tr><td>50 bis 59</td><td>4750</td></tr><tr><td>60 bis 69</td><td>5700</td></tr><tr><td>70 bis 79</td><td>6650</td></tr><tr><td>80 bis 89</td><td>7600</td></tr><tr><td>90</td><td>8550</td></tr></table></td><td></td><td></td></tr>
<tr><td>NoTherapeuticAnticoagulationWCertoparin</td><td>drugAdministration</td><td>$atcde#B01AB13 "Certoparin"<br><br>$sct#395961003 "Certoparin (substance)"</td><td>subcutaneous</td><td>8000 IE 2x/d</td><td></td><td></td></tr>
<tr><td rowspan="2">NoTherapeuticAnticoagulationWUFHOrArgatra</td><td></td><td>/: #any-of</td><td></td><td>CAVE: aPTT values here!</td><td></td><td></td></tr>
<tr><td>drugAdministration</td><td>$atcde#B01AB01 "Heparin"<br><br>$sct#372877000 "Heparin (substance)"</td><td>intravenous</td><td>$loinc#3173-2 "aPTT in Blood by Coagulation assay"</td><td>aPTT >= 50 s</td><td></td></tr>
</table>

<table>
<tr><td></td><td></td><td></td><td>drugAdministration</td><td>$atcde#B01AE03 "Argatroban"<br><br>$sct#116508003 "Argatroban (substance)"</td><td>intravenous</td><td></td><td>aPTT >= 50 s</td><td></td></tr>
<tr><td></td><td></td><td>NoTherapeuticAnticoagulationWTinzaparin</td><td>drugAdministration</td><td>$atcde#B01AB10 "Tinzaparin"<br><br>$sct#412608008 "Tinzaparin (substance)"</td><td>subcutaneous</td><td>175 IE/kg 1x/d</td><td></td><td></td></tr>
<tr><td></td><td></td><td>TherapeuticAnticoagulationFondaparinux</td><td>drugAdministration</td><td>$atcde#B01AX05 "Fondaparinux"<br><br>$sct#708189008 "Fondaparinux (substance)"</td><td>subcutaneous</td><td><table><tr><th>Weight (kg)</th><th>Dose (1x /d)</th></tr><tr><td>< 50</td><td>5 mg</td></tr><tr><td>>= 50, <= 100</td><td>7.5 mg</td></tr><tr><td>> 100</td><td>10 mg</td></tr></table></td><td></td><td></td></tr>
</table>

### 35 - Tidal volume (English)

#### Recommendation

##### Guideline

S3 guideline Recommendations for the inpatient treatment of patients with COVID-19 – Living Guideline - https://register.awmf.org/de/leitlinien/detail/113-001LG

##### Summary

###### English (machine translation)

In ventilated patients with COVID-19 and ARDS, tidal volume should be  6 ml/kg standard body weight, end-inspiratory airway pressure  30 cm H2O.

##### Justification

###### English (machine translation)

At the onset of the COVID-19 pandemic, various editorials and smaller case series suggested that COVID-19 ARDS was atypical because, at least in a subset of cases, it differed from "classic ARDS" in the early phase by higher compliance, reduced recruitable, and high shunt fraction (145, 258). However, in the most recently published larger studies, there were no significant differences in lung compliance, ventilatory pressures, and pressures in patients with COVID-19 associated ARDS in the later course compared with other causes of ARDS (259-261). Therefore, because of the lack of randomized trials of ventilatory therapy in COVID-19, recommendations for ventilatory therapy are derived from the most recently published guidelines on invasive ventilation for acute respiratory failure (193, 195). This includes the recommendations on tidal volume ( 6 ml/kg ideal body weight) and end-inspiratory airway pressure (PEI)  30 cm H2O).

#### Population

| **Name** | **Description** | **Criteria** | | | | |
|---|---|---|---|---|---|---|
| | | **Inclusion / Exclusion** | **Name** | **Category** | **definition.type** | **definition.value** |
| VentilatedCOVID19patientsWithARDS | Ventilated COVID-19 patients with ARDS | | COVID-19 | Condition | SCT 404684003 "Clinical finding (finding)" | $sct#840539006 "Disease caused by Severe acute respiratory syndrome coronavirus 2 (disorder)" |
| | | | Ventilated | Procedure | SCT 71388002 "Procedure (procedure)" | $sct#40617009 "Artificial respiration (procedure)" |
| | | | ARDS | Condition | SCT 404684003 "Clinical finding (finding)" | $sct#67782005 "Acute respiratory distress syndrome (disorder)" |

#### Intervention

The used LOINC code for tidal volume uses "per body weight" but it should use "per *ideal*  body weight" – however, we couldn't find a code for that (neither in LOINC nor in SNOMED CT)

| **Name** | **Description** | **Population** | **Action/Activities** | | | |
|---|---|---|---|---|---|---|
| | | | **Name** | **Action Categroy** | **Goal target measure** | **Goal target detail** |

| | | | | | | |
|---|---|---|---|---|---|---|
| VentilatedCOVID19PatientsWithARDSVentilationPlan | Manage ventilation such that tidal volume 6 ml/kg standard body weight, end-inspiratory airway pressure 30 cm H2O. | VentilatedCOVID19patientsWithARDS | Tidal Volume | ventilatorManagement | $loinc#20117-8 "Tidal volume.spontaneous+mechanical/Body weight [Volume/mass] --on ventilator" | range.high = 6 'ml/kg' "ml/kg" |
| | | | End-inspiratory airway pressure | ventilatorManagement | code changed<br><br>$loinc#76259-1 "Pressure.plateau Respiratory system airway --on ventilator" | range.high = 30 'cm [H2O]' "cm [H2O]" |

### 36a - PEEP (English)

#### Recommendation

##### Guideline

S3 guideline Recommendations for the inpatient treatment of patients with COVID-19 – Living Guideline - https://register.awmf.org/de/leitlinien/detail/113-001LG

##### Summary

###### English (machine translation)

For the orientational adjustment of PEEP in COVID-19, the FiO2/PEEP table of the ARDS Network should be considered. Close monitoring can adjust the PEEP to the individual patient's situation.

##### Justification

###### English (machine translation)

Regarding the setting of positive end-expiratory pressure (PEEP), for patients in the early phase (without classic consolidations, high compliance, expected low recruitable) the PEEP setting according to the values of the LOW-FiO2/PEEP table seems reasonable. In the classic image-morphologic manifestation of ARDS with reduced compliance, the setting should rather be according to the High FiO2/PEEP table (195, 225).
In ARDS and a PaO2/FiO2 < 150 mmHg, abdominal positioning should be consistently performed, with an abdominal positioning interval of at least 16 hours (195). In individual cases, application of inhaled NO, muscle relaxation, or a recruitment maneuver may be considered to bridge severe hypoxemia.
In patients with severe ARDS and refractory hypoxemia (PaO2/FiO2 quotient < 80 or 60 mmHg), the use of venovenous ECMO is a therapeutic option to stabilize gas exchange. However, ECMO should only be considered if all other therapeutic measures have been exhausted, no contraindications exist, and the patient's will has been evaluated in this regard. In a recent French study, the mortality of COVID-19 patients treated with venovenous ECMO was 54% at 90 days (262).
Management of analgesia and sedation in ICU-treated patients should be targeted and monitored using validated measurement tools (263). At most, the sedation goal includes mild sedation, especially with regard to the side effects of sedatives such as Delirium, respiratory depression, hypotension, and immunosuppression. Deep sedation and oversedation are also risk factors for poorer outcomes in COVID-19 sufferers.

#### Population

- For each FiO2 value (0.3, 0.4 etc) we have defined a different population
- The selection works via "valueRange", i.e. the range datatype, with valueRange.low and valueRange.high
- According to the FHIR specicfication for the Range datatype, these values (low/high) are inclusive, and there is no possibility to change that behaviour (because low and high use the SimpleQuantity datatype, which disallows the use of the comparator field, which could have  been used otherwise to specify inclusive or exclusive)
- There is apparently no real way to define an exact cover using the Range datatype in FHIR

| Name | Description | Criteria | | | | |
|---|---|---|---|---|---|---|
| | | Inclusion + / Exclusion − | Name | Type | definition.type | definition.value |
| PopulationVentilatedCOVID19patientsWithARDS-fio2-[0.3, 0.4, 0.5, 0.6, 0.7, 0.8, 0.9] | Ventilated Patients suffering from COVID-19 induced ARDS | + | COVID-19 | Condition | SCT 404684003 "Clinical finding (finding)" | $sct#840539006 "Disease caused by severe acute respiratory syndrome coronavirus 2 (disorder)" |
| | | + | ventilated | Procedure | SCT 71388002 "Procedure (procedure)" | $sct#40617009 "Artificial respiration (procedure)" |

| | | | | | | |
|---|---|---|---|---|---|---|
| | | | FiO2 | ventilationObservableLOINC | $loinc#3150-0 "Inhaled oxygen concentration"<br>• as used in GECCO FiO2<br>• Alternative (as decimal, without % unit): 71835-3 Oxygen/Gas total [Pure volume fraction] Inhaled gas | FiO2 [0.0 0.4, 0.5, 0.6, 0.7, 0.8, 0.9] |

### Intervention

| Name | Description | Criteria | | | |
|---|---|---|---|---|---|
| | | **Name (Goal ID)** | **Action Category** | **Goal target measure** | **Goal target detail** |
| VentilatedCOVID19patientsWithARDSInterventionPlan-fio2-[0.0, 0.4, 0.5, 0.6, 0.7, 0.8, 0.9] | PEEP Intervention Plan for Ventilated COVID-19 Patients with ARDS, inspiratory oxygen fraction currently -[0.0, 0.4, 0.5, 0.6, 0.7, 0.8, 0.9] | ventilator-management-fio2-0.0-goal | ventilatorManagement | $loinc#76248-4 "PEEP Respiratory system --on ventilator" | >=5 cm [H2O] |
| | | ventilator-management-fio2-0.4-goal | ventilatorManagement | $loinc#76248-4 "PEEP Respiratory system --on ventilator" | >=5 cm [H2O] |
| | | ventilator-management-fio2-0.5-goal | ventilatorManagement | $loinc#76248-4 "PEEP Respiratory system --on ventilator" | >=8 cm [H2O] |
| | | ventilator-management-fio2-0.6-goal | ventilatorManagement | $loinc#76248-4 "PEEP Respiratory system --on ventilator" | >=10 cm [H2O] |
| | | ventilator-management-fio2-0.7-goal | ventilatorManagement | $loinc#76248-4 "PEEP Respiratory system --on ventilator" | >=10 cm [H2O] |
| | | ventilator-management-fio2-0.8-goal | ventilatorManagement | $loinc#76248-4 "PEEP Respiratory system --on ventilator" | >=14 cm [H2O] |
| | | ventilator-management-fio2-0.9-goal | ventilatorManagement | $loinc#76248-4 "PEEP Respiratory system --on ventilator" | >=14 cm [H2O] |
| | | ventilator-management-fio2-1.0-goal | ventilatorManagement | $loinc#76248-4 "PEEP Respiratory system --on ventilator" | >=18 cm [H2O] |

### Notes

Since the recommendation is ambiguous (who is considered to be an early-phase ARDS patient?, what is "well ventilated/oxygenated"? etc.), we have decided to model the following: For a patient with a given FiO2, is the PEEP-value greater or equal than the minimum level specified for this FiO2. Reference is the table "Lower PEEP, higher fiO2" from the AWMF Recommendation for ARDS (https://www.awmf.org/fileadmin/user_upload/Leitlinien/001_Anaesthesiologie_und_Intensivmedizin/001-021kt_S3_Invasive_Beatmung_2017-12.pdf)

| FiO2 | 0.0 | 0.3 | 0.4 | 0.5 | 0.6 | 0.7 | 0.8 | 0.9 | 1.0 |
|---|---|---|---|---|---|---|---|---|---|
| min. PEEP | 5 | 5 | 5 | 8 | 10 | 10 | 14 | 14 | 18 |

The FiO2 0.0 with PEEP = 5 has been added by us and is not part of the ARDS network table

### 36b - Prone position (English)

#### Recommendation

##### Guideline

S3 guideline Recommendations for the inpatient treatment of patients with COVID-19 – Living Guideline - https://register.awmf.org/de/leitlinien/detail/113-001LG

##### Summary

###### English (machine translation)

In ARDS and a PaO2/FiO2 < 150 mmHg, abdominal positioning should be consistently performed, with an abdominal positioning interval of at least 16 hours.

##### Justification

###### English (machine translation)

Regarding the setting of positive end-expiratory pressure (PEEP), for patients in the early phase (without classic consolidations, high compliance, expected low recruitable) the PEEP setting according to the values of the LOW-FiO2/PEEP table seems reasonable. In the classic image-morphologic manifestation of ARDS with reduced compliance, the setting should rather be according to the High FiO2/PEEP table (195, 225).
In ARDS and a PaO2/FiO2 < 150 mmHg, abdominal positioning should be consistently performed, with an abdominal positioning interval of at least 16 hours (195). In individual cases, application of inhaled NO, muscle relaxation, or a recruitment maneuver may be considered to bridge severe hypoxemia.
In patients with severe ARDS and refractory hypoxemia (PaO2/FiO2 quotient < 80 or 60 mmHg), the use of venovenous ECMO is a therapeutic option to stabilize gas exchange. However, ECMO should only be considered if all other therapeutic measures have been exhausted, no contraindications exist, and the patient's will has been evaluated in this regard. In a recent French study, the mortality of COVID-19 patients treated with venovenous ECMO was 54% at 90 days (262).
Management of analgesia and sedation in ICU-treated patients should be targeted and monitored using validated measurement tools (263). At most, the sedation goal includes mild sedation, especially with regard to the side effects of sedatives such as Delirium, respiratory depression, hypotension, and immunosuppression. Deep sedation and oversedation are also risk factors for poorer outcomes in COVID-19 sufferers.

#### Population

| Name | Description | Criteria | | | | |
|---|---|---|---|---|---|---|
| | | **Inclusion + / Exclusion −** | **Name** | **Category** | **definition. type** | **definition.value** |
| VentilatedCOVID19patientsWithARDSWithOxygenationFailure | Ventilated Patients suffering from COVID-19 induced ARDS which exhibit oxygenation failure | + | COVID-19 | Condition | SCT 404684003 "Clinical finding (finding)" | $sct#840539006 "Disease caused by severe acute respiratory syndrome coronavirus 2 (disorder)" |
| | | + | Ventilated | Procedure | SCT 71388002 "Procedure (procedure)" | $sct#40617009 "Artificial respiration (procedure)" |
| | | + | ARDS | Condition | SCT 404684003 "Clinical finding (finding)" | $sct#67782005 "Acute respiratory distress syndrome (disorder)" |
| | | + | hypoxic respiratory failure | laboratory | $loinc#50984-4 " Horowitz index in Arterial blood" | Horowitz (=oxygenation) index lower than 150 mmHg |

#### Intervention

| Name | Description | Population | Action/Activities | | | |
|---|---|---|---|---|---|---|
| | | | **Name** | **Type** | **Action.code** | **Timing** |

| | | | | | | |
|---|---|---|---|---|---|---|
| PronePositioning | Actively putting the patient in prone position, for at least 16 hours at a time | VentilatedCOVID19patientsWithARDSWithOxygenationFailure | prone positioning | bodyPositioning | $sct#431182000 "Placing subject in prone position (procedure)" | >= 16h |

### Sepsis M.1 - Tidal volume (English)

#### Recommendation

##### Guideline

Sepsis - prevention, diagnosis, treatment and aftercare - https://www.awmf.org/leitlinien/detail/ll/079-001.html

##### Summary

###### English (machine translation)

We recommend ventilating patients with ARDS with a VT  6 ml/kg standard body weight (bw). (Table 1, Appendix)

##### Justification

###### English (machine translation)

Several meta-analyses of RCTs comparing ventilation with small VT or low end-inspiratory airway pressure (PEI) < 30 cm H2O (resulting in VT < 7 ml/kg bw) versus ventilation with VT 10 to 15 ml/kg bw with and without change in positive end-expiratory pressure (PEEP) show a reduction in mortality in adult (> 16 years) invasively ventilated patients with ARDS. The current SSC international guideline recommends for ventilation in patients with ARDS presenting with more severe sepsis or septic shock a VT  6 ml/kg standard CG (strong recommendation, high quality evidence) to aim for a PEI < 30 cm H2O (strong recommendation, moderate quality evidence).

Based on this demonstrated reduction in mortality in patients with ARDS by ventilation with low tidal volumes in the absence of evidence of relevant harm, the Guideline Committee assigns a strong recommendation for ventilation of invasively ventilated patients with ARDS with a tidal volume VT  6 ml/kg standard CG. The use of VT  6 ml/kg standard CG to achieve a PEI < 30 cm H2O may result in varying degrees of hypercapnia and respiratory acidosis. The resulting hypercapnia and respiratory acidosis were tolerated and treated to varying degrees in the RCTs. Hypercapnia and respiratory acidosis can increase intracranial pressure, exacerbate pulmonary hypertension and myocardial depression, and reduce renal blood flow in critically ill patients. Therefore, in patients in whom hypercapnia and respiratory acidosis should be avoided because of the underlying disease, the benefits and risks of reducing VT should be evaluated on an individual basis.

#### Population

> ⓘ The recommendation has been selected by the clinicians based *only* on the summary text and therefore, sepsis/septic shock patients are not included as population criteria.

| **Name** | **Description** | **Criteria** | | | | |
|---|---|---|---|---|---|---|
| | | **Inclusion ⊕ / Exclusion ⊖** | **Name** | **Category** | **definition.type** | **definition.value** |
| PopulationVentilatedARDSPatients | Ventilated Patients with ARDS | ⊕ | ARDS | Condition | SCT 404684003 "Clinical finding (finding)" | $sct#67782005 "Acute respiratory distress syndrome (disorder)" |
| | | ⊕ | Ventilated | Procedure | SCT 71388002 "Procedure (procedure)" | $sct#40617009 "Artificial respiration (procedure)" |

#### Intervention

> ⓘ The used LOINC code for tidal volume uses "per body weight" but it should use "per *ideal* body weight" – however, we couldn't find a code for that (neither in LOINC nor in SNOMED CT)

| Name | Description | Population | Action/Activities | | | |
|---|---|---|---|---|---|---|
| | | | **Name** | **Action Category** | **Goal target measure** | **Goal target detail** |
| VentilatedARDSPatients InterventionPlan | Manage ventilation such that VT 6 ml/kg standard body weight (bw). | PopulationVentilatedARDSPatients | Tidal Volume | ventilatorManagement | CODEX-CELIDA CodeSystem:<br><br>#tvpibw "Tidal volume / ideal body weight (ARDSnet)" | range.high = 6 'ml/kg' "ml/kg" |
